# Barriers and facilitators to implementing synchronous telehealth interventions for people with dementia - a systematic review

**DOI:** 10.1101/2025.09.30.25336993

**Authors:** Richard Talbot, Charlotte Harris, Esther Cable, Breege Whiten, Jason D. Warren, Rosemary Varley, Anna Volkmer

**Author notes:** Corresponding author: Richard Talbot 202 Chandler House, 2 Wakefield Street, London WC1N 1PF. @rtslt, @profjasonwarren, @volkmer_anna.

## Abstract

**Introduction:** Behavioural interventions, such as those provided by allied health professionals and psychologists, help manage symptoms of people living with dementia. Access to such interventions depends on individual factors (e.g., support, travel) and service availability. One way to improve access and availability is via synchronous telehealth. However, there may be challenges implementing telehealth interventions.

**Method:** A systematic review was conducted to explore barrier and facilitators to implementing synchronous telehealth intereventions for people living with dementia. The review protocol was registered on PROSPERO (CRD42024508098). Five databases were systematically searched in February 2024. Implementation determinants were extracted and analysed deductively using the Theoretical Domains Framework (TDF) and COM-B behaviour change models. Determinants that did not correspond with the TDF or COM-B were described inductively using content analysis. Determinants were further analysed as to whether they pertained to the intervention, or to the technology used to deliver it.

**Results:** 25 papers were included in the review, including group, dyadic and individual interventions from 14 countries including those from the Global South. Key implementation determinants corresponded to the TDF domains *Environmental Context & Resources and Knowledge*. Two inductively created domains were developed: *Creativity & Safety*. Most implementation barriers were to do with technology. These were often clear problems with potentially straightforward, linear, implementation solutions related to the COM-B concepts of *Capability* and *Opportunity*. Most implementation facilitators involved the telehealth intervention itself. These were often complex issues, with potentially complex and non-linear implementation strategies, most often related to the concept of *Motivation*. Reporting quality was variable in the literature.

**Conclusion:** Use of implementation and behaviour change frameworks may enhance reporting, reproducibility, and improve implementation. Such frameworks may also help prioritise determinants to address and delineate potential implementation strategies.

## Background

Dementia describes a group of neurodegenerative diseases which progressively and differentially impact multiple neurocognitive domains including memory, motor function, behaviour and language (World Health Organisation, 2025). In 2021, approximately 57 million people worldwide were estimated to be living with dementia, with 10 million new cases diagnosed annually. Over 60% of these cases are predicted to come from in low- and middlelZlincome countries (World Health Organisation, 2025). By 2050, the total number of people living with dementia is set to reach over 152 million (Alzheimer’s Disease International, 2017). With recent pharmaceutical advances in dementia treatment (e.g., Hardy & Mummery, 2023; Mummery et al., 2023), behavioural interventions (i.e., non-pharmacological) for dementia symptom management - such as those provided through speech and language therapy (SLT), physiotherapy (PT), occupational therapy (OT), psychology, care and caregiver support - are also showing positive impacts for people with dementia and their care partners (Morello et al., 2017; Woods et al., 2023; Livingston et al., 2024). However, access to such interventions is impacted by individual circumstances including travel constraints and availability of a support network (Gulline et al., 2025). Service availability is also variable across healthcare providers, settings, and geography, particularly for rarer dementias (Hockley et al., 2025; Sullivan et al., 2025).

One way to increase the reach and availability of behavioural interventions for dementia is via synchronous telehealth, i.e., intervention delivered synchronously online via videoconferencing or telephone. Telehealth delivery may increase access to interventions, enhancing the ability of people living with dementia - and their families and friends - to engage with and benefit from behavioural interventions. This may be particularly relevant for those unable to attend clinic-based sessions, such as people in the later stages of the disease. It also enables care partners with work or caring commitments to attend or support sessions (e.g., Volkmer et al., 2023a). As such, telehealth delivery may influence both the implementation and uptake of the interventions.

The COVID-19 pandemic necessitated a rapid and substantial expansion of telehealth usage (Omboni et al., 2022; Shaver, 2022). While adoption and attitudes during this period may not be typical, the pandemic has likely permanently reshaped the role and perception of telehealth. If the usage of telehealth remains more widespread, it will be important to identify determinants to better implement and sustain telehealth intereventions (Thomas et al., 2024).

### Implementation science, complexity, and telehealth in dementia

Implementation science is “the study of methods and strategies to promote the adoption and integration of proven clinical treatments, practices, organisational, and/or management interventions into routine practice.” (Neuman et al.. 2020, p 434). While effectiveness research focusses on how interventions work in real-world settings, implementation science focusses on ways to enable the “spread and adoption”, integration and sustainability of evidence-based interventions in real-world settings (Glasgow et al., 2012: p. 1275).

Delays in translating health research into clinical practice result in reduced patient access to evidence-based interventions and wasted research and development effort (Morris et al., 2011). Over half of clinical innovations never make it into general clinical usage (Bauer & Kirchner, 2020). Furthermore, interventions which are poorly implemented in everyday practice may not produce the same health benefits seen in research trials, leading to poor clinical outcomes (Neuman et al., 2020). Considering the pace of technological advance, implementation of telehealth interventions may be particularly problematic. To address the research-practice gap, the Medical Research Council (MRC) framework for evaluating complex interventions emphasises that implementation should be considered early and throughout intervention development to increase the chance of timely, widespread and sustained adoption of interventions. Implementation questions should be addressed at each stage of intervention development and evaluation (Skivington et al., 2021). Implementation processes are therefore iterative. Implementation questions go beyond mere questions of effectiveness to consider how implementable interventions will be across dynamic and evolving contexts such as service settings, individuals’ circumstance, or socio-cultural norms (Raine et al., 2016). This may be especially relevant in dementia (where symptoms evolve over time and interact with individuals’ lives and overall quality of lie), telehealth (evolving technology and use within organisations), and behavioural interventions (changing therapeutic protocols and relationships over time).

Implementation processes must be both non-linear and consider complexity. The Cynefin framework (Snowden, 2002; Snowden et al., 2022) has been used in healthcare and implementation science (e.g., Lunghi & Baroni, 2019) to help identify problems as *clear*, *complicated* or *complex* (see Box 1).

#### Box 1

##### Cynefin framework overview: simple, complicated, and complex problems

- **Clear** - Clear cause-and-effect, well-established processes exist, with potentially straightforward solutions. Outcomes are predictable.
- **Complicated** - Cause-and-effect relationships exist but require expertise or analysis to identify the best solution and solve unforeseen factors. Outcomes are predictable.
- **Complex** - Cause-and-effect relationships are unclear, solutions emerge through experimentation and learning over time. Behaviours emerge through interaction between people and systems. Outcomes are unpredictable.

Understanding the type of problem increases the likelihood of identifying potential solutions. In healthcare management and implementation, experts have been criticised for mischaracterising complex problems as complicated ones, leading to inappropriate solutions that overlook the complexity, while concurrently overlooking clear problems that could more straightforwardly be addressed (Cottam, 2018; Glouberman & Zimmerman, 2002; Haynes & Loblay, 2024).

Using theoretical frameworks to systematically identify implementation determinants (i.e., barriers and facilitators) can lead to more effective implementation strategies, and better understanding of implementation outcomes (Eccles et al., 2005; Haynes & Loblay, 2024). The Theoretical Domains Framework (TDF) (Cane et al., 2012) is a comprehensive, integrative, meta-theoretical framework which helps to identify behaviour change factors across fourteen domains and thereby investigate implementation determinants (factors influencing implementation) and strategies (ways to leverage facilitators and overcome barriers and thus improve outcomes) (Atkins et al. 2017; Phillips et al., 2015). The TDF has been informative in understanding behaviour change and implementation in healthcare interventions (Cane et al., 2012; Cowdell & Dyson, 2019; Schrubsole et al., 2023).

However, some limitations of the of the TDF have been described including lack of domain specificity (Atkins et al., 2017), insufficient specification of how domains may interact (Cowdell & Dyson, 2019; Lipworth et al., 2013), and difficulty accounting for context, such as socio-cultural factors impacting behaviour and implementation (Mather et al., 2022). Thus, the framework has been enhanced using various methods. For instance, researchers have applied the TDF using a deductive approach (aligning data within the TDF domains) alongside inductive reasoning. This enables categorisation of content that does not neatly fit within the existing TDF domains. This approach ensures that important but unexpected themes are not overlooked,. As an example, the TDF has been combined with content analysis (Elo & Kyngäs, 2008; Hsieh & Shannon, 2005) by Schrubsole and colleagues (2023) to identify and then synthesise determinants to speech and language therapists implementing communication partner training (Schrubsole et al., 2023). The TDF has also been used alongside reflexive thematic analysis (Braun & Clarke, 2006) to better account the impact of socio-cultural factors on complex healthcare interventions (e.g., Nguyen-Hoang et al., 2024; Petersen et al., 2023).

To overcome difficulties in identifying how TDF domains may combine in complex ways, the TDF has been integrated with complementary frameworks such as the COM-B model. This model is structured into three domains (Capability, Opportunity, and Motivation) that have been proposed as pivotal in changing behaviour (Michie et al., 2011, 2014). The COM-B has been used alongside the TDF in healthcare implementation systematic reviews (e.g., Atkins et al., 2020; Mather et al., 2022; Shrubsole et al., 2023) and telehealth implementation research (e.g., MacPherson & Kapadia, 2023). The TDF has also been combined with the Consolidated Framework for Implementation Research (CFIR; Damschroder et al., 2009) to better account for system level determinants alongside determinants focussed on individuals (e.g., O’Donovan et al., 2023; Birken et al., 2017).

### Existing reviews of telehealth use in dementia interventions

No systematic reviews have investigated implementation determinants of telehealth for people with dementia which included all dementia types and used a theoretical framework to analyse and synthesise the data. A systematic review by Yi et al. (2021) on telehealth use in dementia care focused only on cognitive assessment and delivery of services (not specific interventions) to people with Alzheimer’s Disease (AD) and Mild Cognitive Impairment (MCI) (Yi et al., 2021). It also emphasised cognitive, sensory, and technological factors, rather than wider implementation factors within a theoretical framework. Adam’s et al (2020) focused on implementation but only included studies with people with AD and Parkinson’s disease, finding that tele-rehabilitation was acceptable and feasible. They reported improved access to specialist care and reduced burden (e.g., travel time) as facilitators. However, they emphasised technical and access barriers due to cognitive change and digital poverty. Liang and Aranda’s (2023) scoping review investigated dyadic telehealth interventions (i.e., including care partners) for people with all types of dementia. This review identified facilitators including the perceived personal and social benefits of telehealth interventions, but again barriers related to digital literacy and poverty. They recommended that future telehealth implementation research in dementia should broaden inclusion by increasing sample diversity to better address digital poverty barriers. More recently, Ye and colleagues (2025) systematically reviewed the literature on implementation factors on telehealth use in dementia. They they also highlighted key themes of digital access barriers, and poor sample diversity, alongside social and personal benefits (Ye et al., 2025). However, both Liang & Aranda and Ye et al. (2025) used no framework to synthesize determinants. Using a simple barrier and facilitators approach in the absence of theory, has been criticised for being overly reductive (Haynes & Loblay, 2024).

Existing reviews therefore either lack a focus on implementation using a theoretical framework (Liang & Aranda, 2023; Ye et al., 2025) or address specific neurodegenerative conditions (Adams, et al., 2020; Yi et al., 2021). This study aims to fill this gap and answer the research question:

What are the barriers and facilitators to implementing behavioural telehealth interventions for people living with dementia, of any type, with or without their care partners.

## Methods

The protocol for this systematic review was pre-registered and published on PROSPERO on the 31^st^ ^of^ January 2024 under registration number CRD42024508098. The protocol follows the PRISMA-P (Moher et al., 2015) and PRISMA E&E (Liberati et al., 2009) transparent reporting of systematic reviews recommendations.

### Search strategy

On the 1^st^ of February 2024, systematic searches were undertaken in five databases: MEDLINE, Embase, PsycINFO (accessed through Ovid) and CINAHL Plus and Scopus; predicted to encompass the dementia intervention literature from medicine, nursing, allied therapies, and psychology.

The search strategy was refined in consultation with research librarians at University College London (co-authors EC and BW). Consequently, the broad terms ‘virtual’ and ‘remote’ were not used to reduce the risk of retrieving papers focusing on non-therapy interventions, e.g., virtual training environments for professionals. The search was limited to articles published since 1^st^ January 2018 because preliminary searches identified an acceleration of dementia telehealth intervention papers published from 2018 onward, including a surge in telehealth use necessitated by the COVID-19 pandemic. Including intervention delivered during the COVID-19 period was predicted to provide evidence on intervention adaptations under pressure, as well as potential lasting shifts in telehealth practice.

The search strategy included the following concepts to search article titles, abstracts and keywords *dementia (and associated terms) AND telehealth (and associated terms) AND intervention (and associated terms)*. See Figure 1 for the full search strategy.

**Figure 1:**
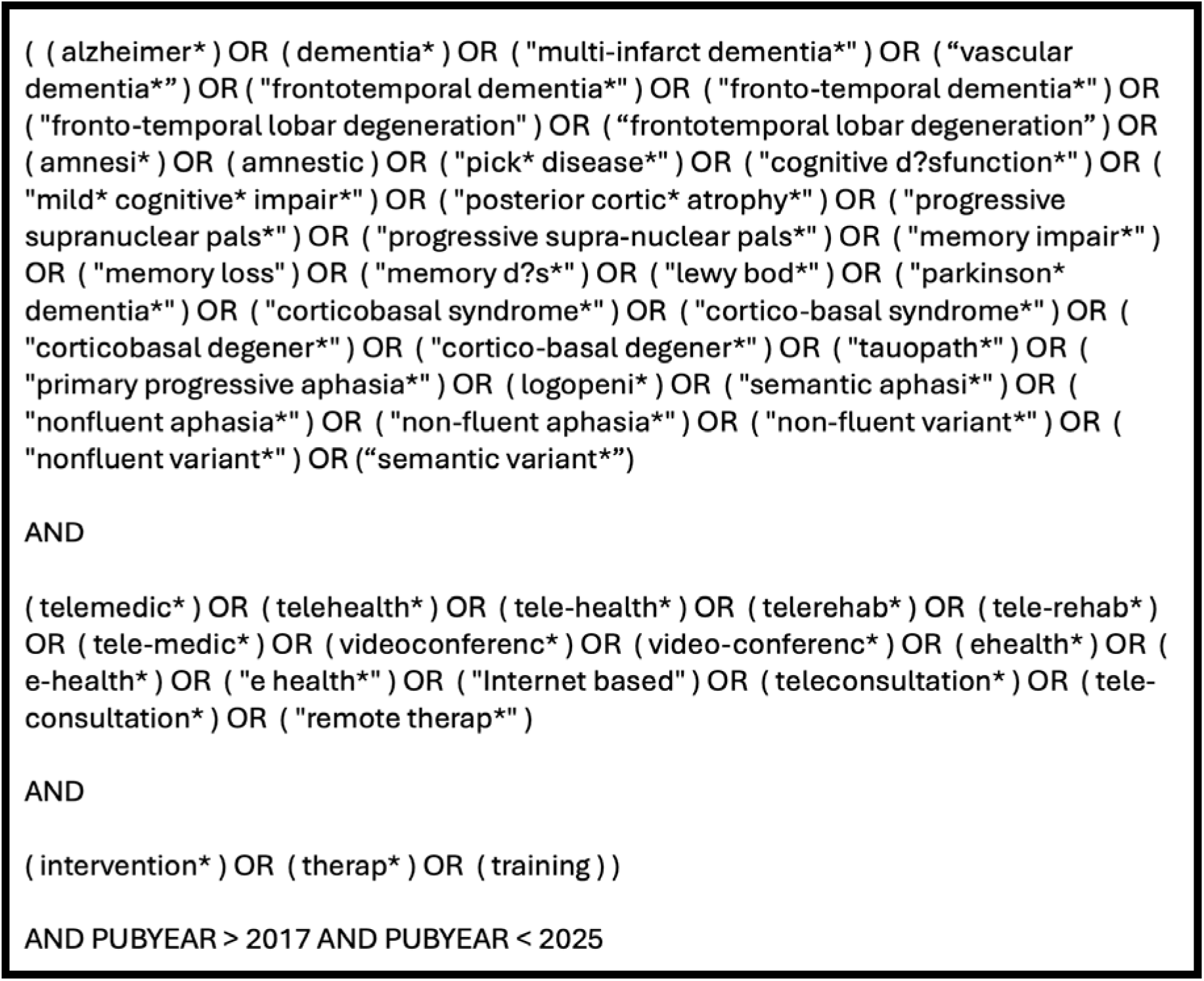
Full search strategy

### Study selection

Co-authors RT and CH independently screened all article titles and abstracts in line with selection criteria (see Table 1). Included full text articles were then independently screened for inclusion by RT and CH. There was 93% agreement for titles/abstracts and 91% for full texts. All discrepancies were resolved through discussion to establish consensus. Reasons for exclusion were documented. Following selection, a snowball method was applied using manual reference chaining and citation tracking of included full-text articles, to identify appropriate papers not included in the original systematic search.

**Table 1:** Selection criteria.

| *Inclusion criteria* |
| --- |
| • describe telehealth (i.e., delivered remotely via videoconferencing or telephone) behavioural, synchronous (i.e. delivered by a therapist or facilitator on-line at the same time as participants) interventions for people with dementia (of any type or disease stage), with or without their caregivers included in the intervention, in any configuration (i.e., group, dyadic, one-to-one or a mixture)<br>• delivered by a health professional, or suitably trained assistant under the supervision of a health professional<br>• original and empirical research studies (i.e., not review papers or opinion pieces)<br>• published in a peer reviewed scholarly journal or academic conference proceedings (to ensure the inclusion of engineering and technology literature).<br>• include adult participants over 18 years of age<br>• report barriers and/or facilitators to the implementation of synchronous remote interventions |
| *Exclusion criteria:* |
| • describe asynchronous interventions (e.g., pharmacological, self-management programs, therapy apps) which are managed independently by participants and do not occur with the necessary presence of a therapist or facilitator<br>• describe an entire service (e.g., a multi-disciplinary gerontology service)<br>• interventions which take place in virtual reality environments<br>• studies which only include those with a diagnosis of MCI, or if it is not possible to distinguish results or analysis relating to clients with dementia from those with an MCI diagnosis |

National Institute of Health Research and Care Excellence (NICE) guidance on ‘Dementia: assessment, management and support for people living with dementia and their carers’ (NICE, 2018) frames dementia intervention as rehabilitation. To align with this, the operational definition of *intervention* used for article inclusion was taken from the World Health Organisation definition of *rehabilitation*, specifically “a set of interventions designed to optimize functioning and reduce disability in individuals with health conditions in interaction with their environment” (World Health Organisation, 2024).

### Quality assurance

RT assessed the quality of papers using the Mixed Methods Appraisal Tool (MMAT) (Hong et al., 2018). The MMAT is a reliable quality assessment instrument, which enables the assessment of qualitative, quantitative and mixed methods study designs within the one tool (Crowe & Sheppard, 2011). Qualitative and quantitative papers are scored on a scale of 0-5. Mixed-methods papers are scored out of 15 by adding qualitative and quantitative scores to a mixed-methods component which is also scored out of 5. No papers were eliminated following quality assessment. Ratings were included to inform the reader of the quality of the studies, and to help describe and understand them.

### Data extraction

Data were extracted from eligible full texts using a Microsoft Excel spreadsheet, structured using the TDF domains. In line with PRISMA-P, co-authors RT and AV independently extracted data from a random >10% sample (n=3; 12%) of papers to calibrate data extraction processes and ensure consistency in applying the *a priori* extraction criteria. Following this, the data extraction spreadsheet was refined (e.g., terms clarified) and RT continued extracting data from the remaining 22 papers.

### Data analysis

Deductive thematic analysis was used to group identified implementation determinants according to behaviour change domains from the TDF (Cane et al., 2012) which were then subsumed within the COM-B model (Michie et al., 2014) using framework analysis (Ritchie & Lewis, 2003). Concepts which did not align with domains in the TDF, but which repeatedly appeared in the retrieved literature, were coded inductively using content analysis and informed the development of new domains (Elo & Kyngäs, 2008; Hsieh & Shannon, 2005). The inductive coding process involved:

- open coding - recording an identified determinant that could not be neatly aligned with the TDF
- categorization - grouping the codes to identify recurring patterns
- abstraction - grouping categorised data into higher order themes

The frequency of deductive and inductive domains was analysed using descriptive statistics and interpreted using narrative synthesis. This process was informed by the system used by Schrubsole et al. (2023). Steps included:

- Summarise the frequency a TDF domain (and corresponding COM-B component) or inductive domain
- Code determinants as to whether they pertained to the intervention, the technology, or a mixture of the two (i.e., it was not possible to disassociate the two concepts)
- Group similar determinants together as exemplar categories of implementation determinants within domains

## Results

The search identified 2,796 articles. Following duplicate removal, 1,297 article titles/abstracts were screened for eligibility. Having excluded 1,251 articles, 46 full texts were retrieved and screened for inclusion. This resulted in the exclusion of a further 23 articles. The references of the remaining 23 articles were then checked against the inclusion and exclusion criteria by RT. This resulted in the in the addition of a further two articles. Thus, a total of 25 articles were included in the review. The PRISMA flow diagram (see Figure 2) provides details of reasons for exclusion.

**Figure 2:**
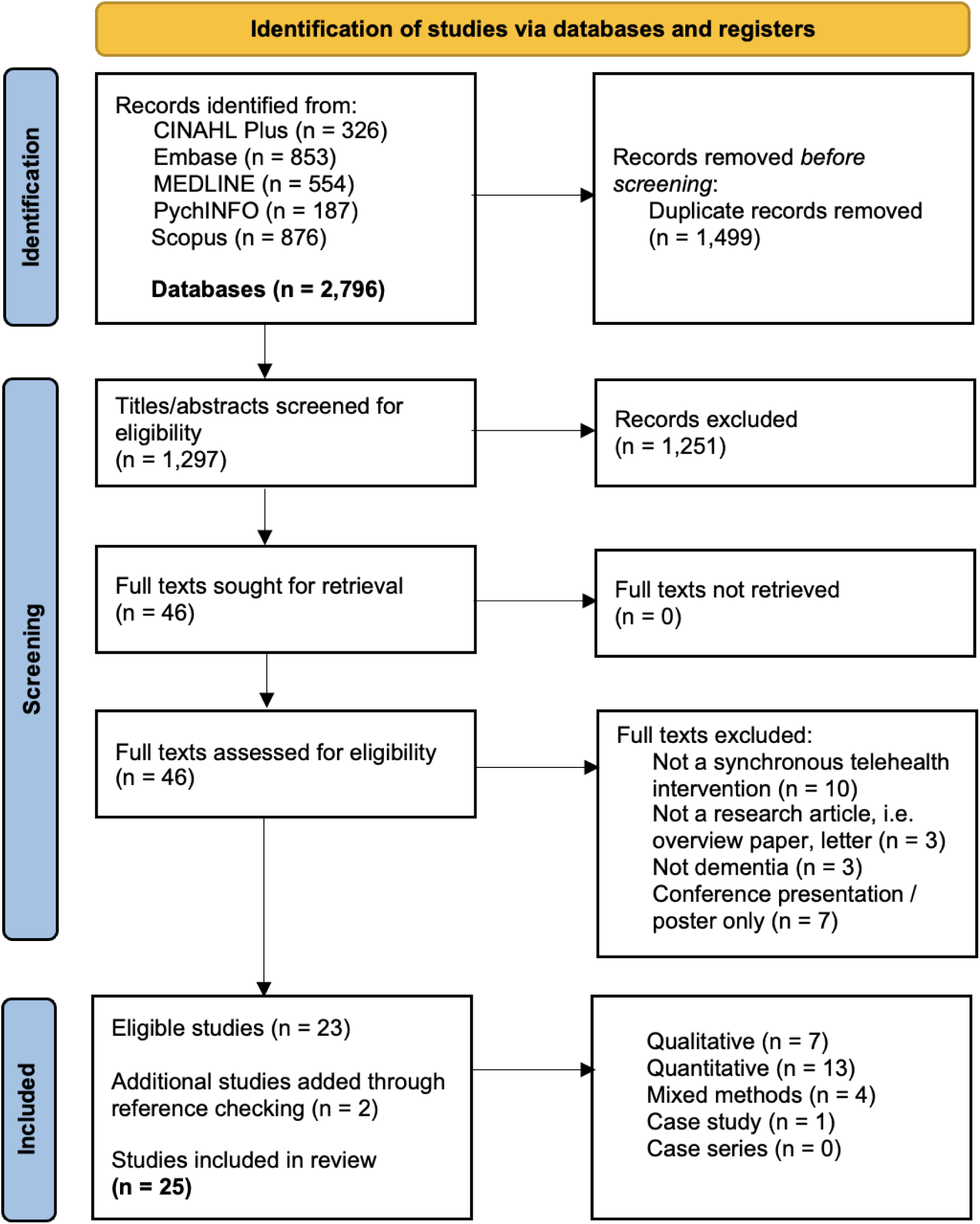
PRISMA flow diagram for selection of studies documenting barriers and facilitators to telehealth implementation in dementia

### Quality assurance

The overall quality of the literature was high. All qualitative papers scored the maximum 5/5. All quantitative papers scored 3/5 or above (five scoring 5/5, six scoring 4/5, and three scoring 3/5). The most frequently absent or unclear items from quantitative studies were ‘Are the confounders accounted for in the design and analysis?’ and ‘During the study period, is the intervention administered (or exposure occurred) as intended?’. The four mixed-methods studies ranged in quality from 9/15 to 15/15 (Average = 12/15). See <u>supplementary file 1</u> for details of the quality assessments.

### Study characteristics

The 25 included papers represented research from 14 countries including the Global South. They included a total of 593 research participants (317 people with dementia, 236 care partners and 40 healthcare professionals). See Table 2 for a summary of characteristics of included studies.

**Table 2:**
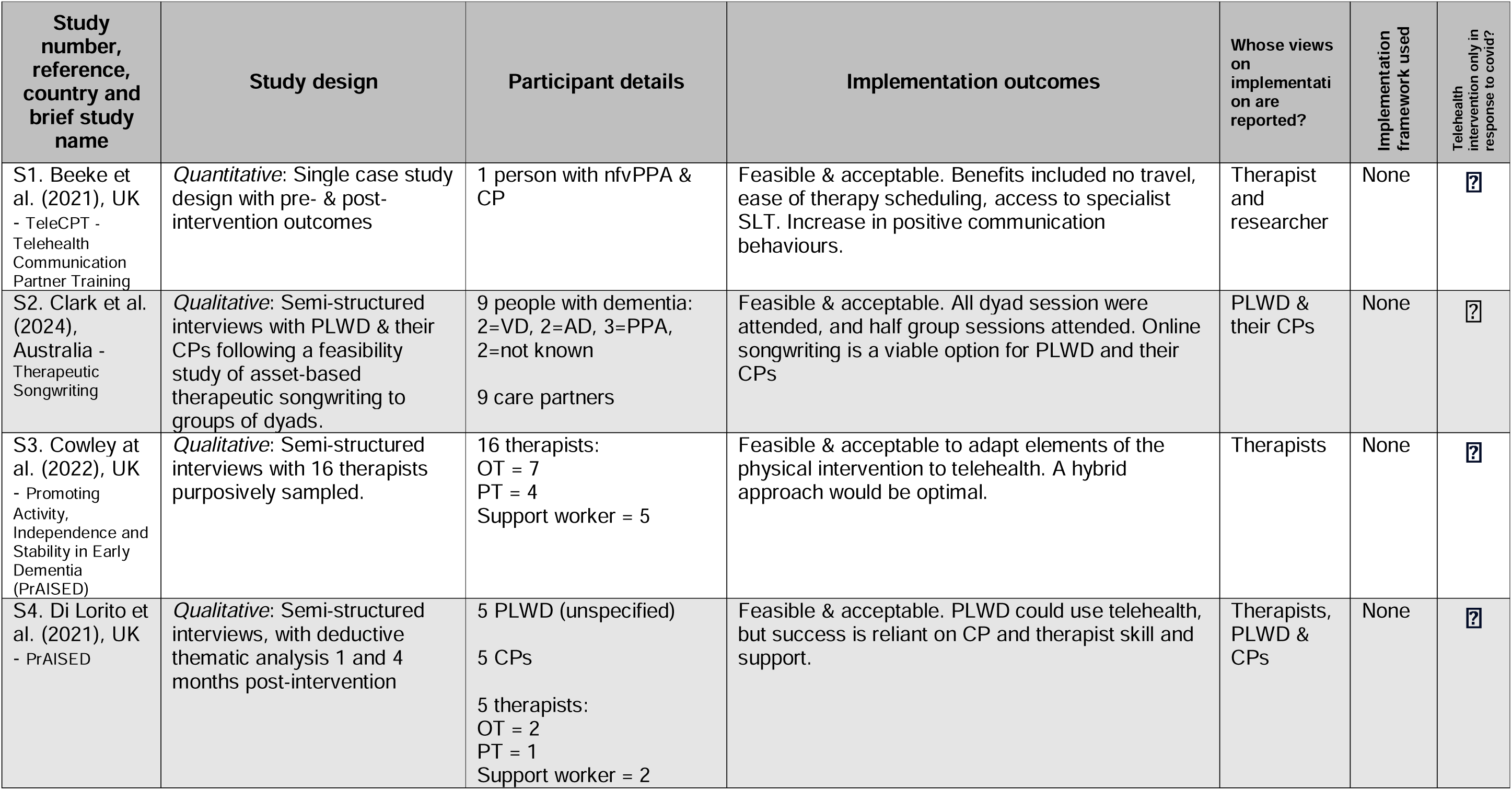
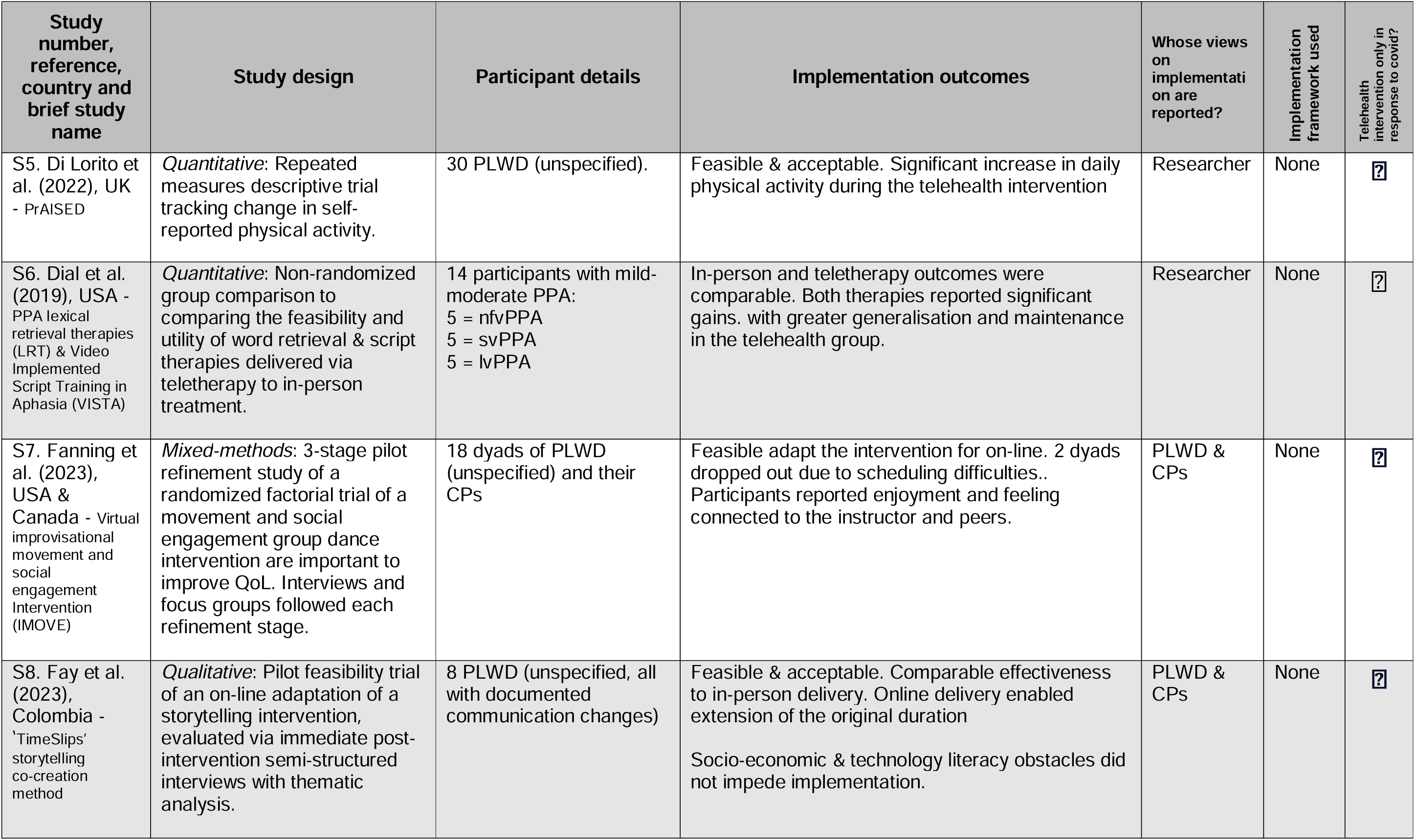
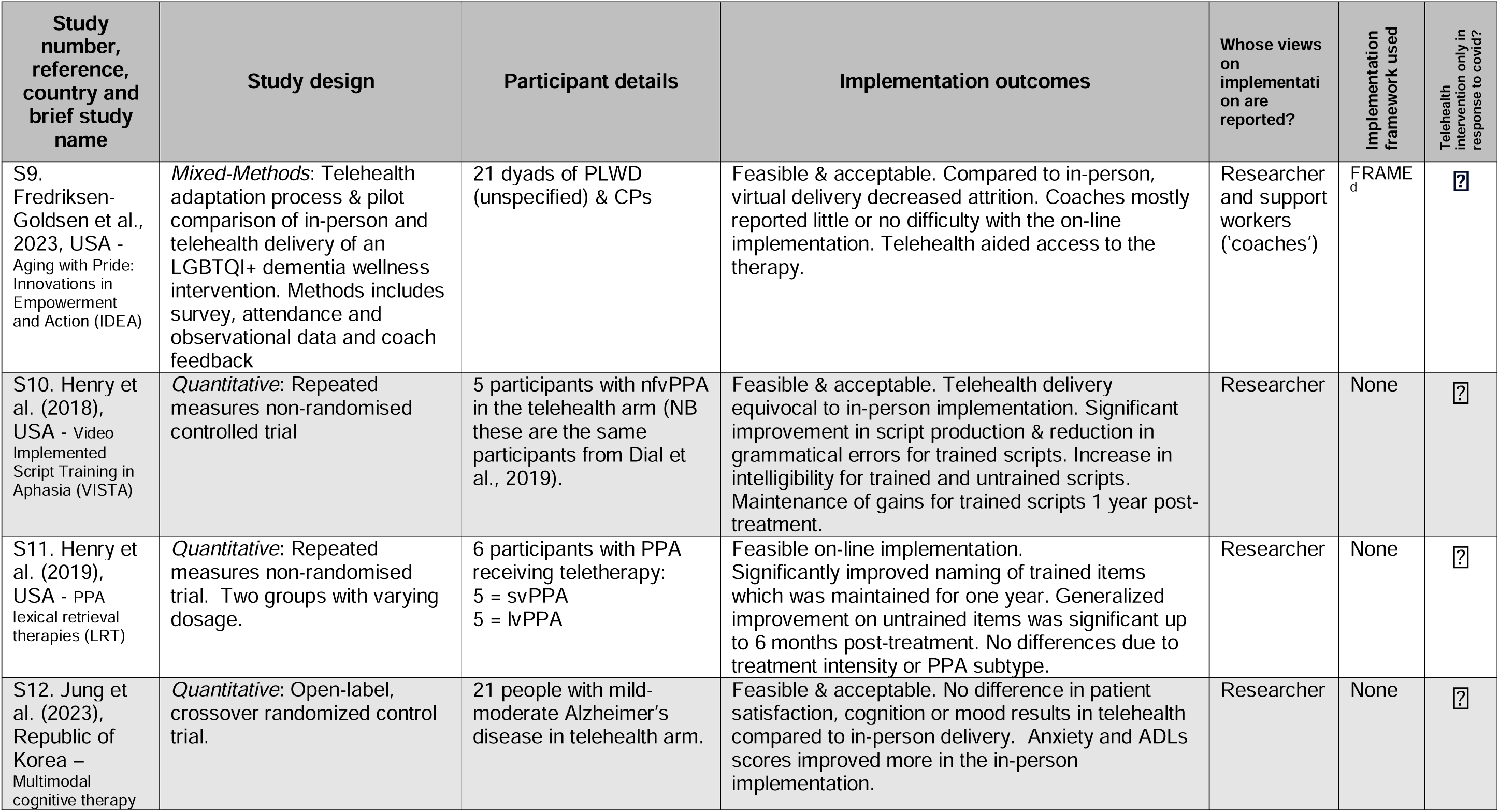
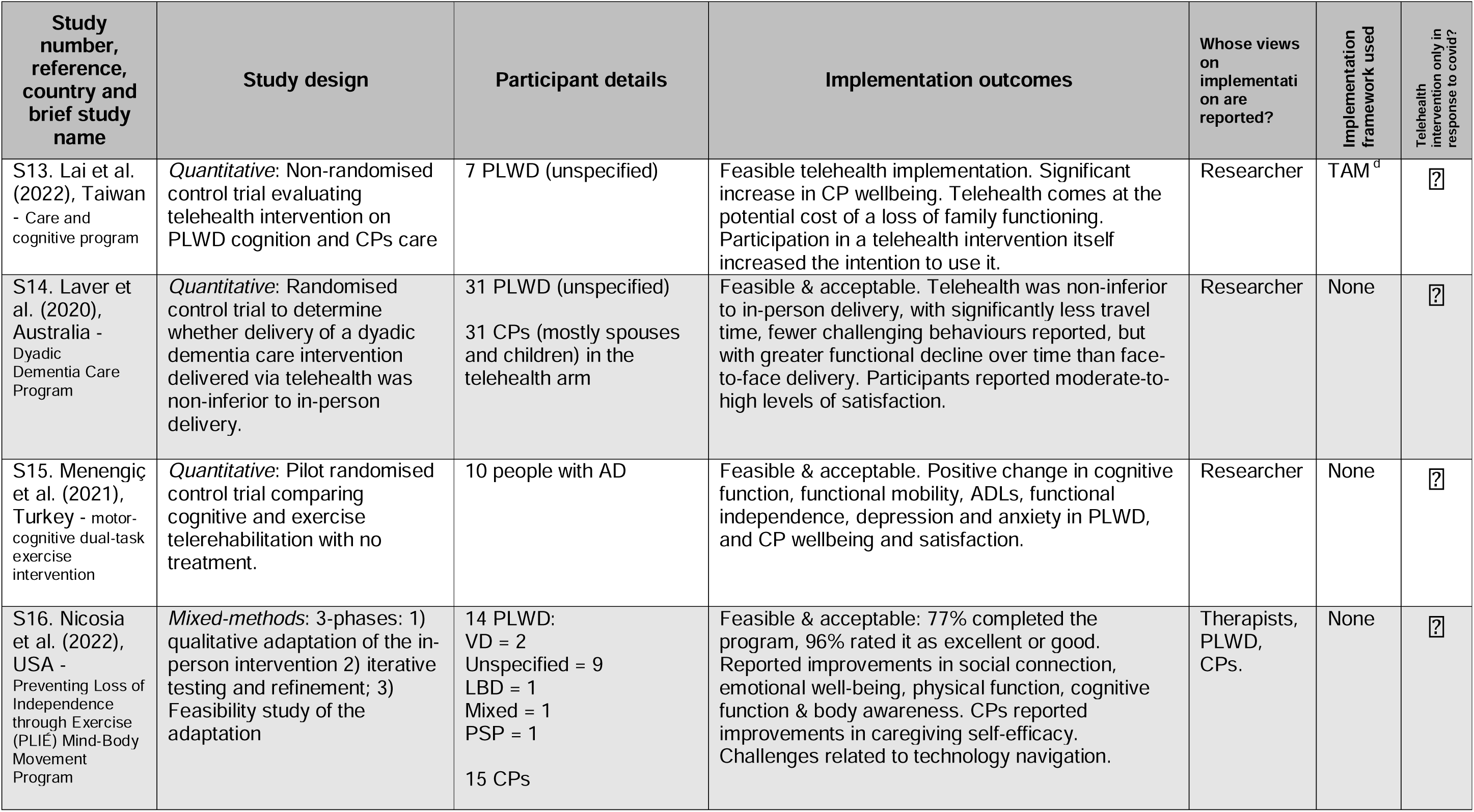
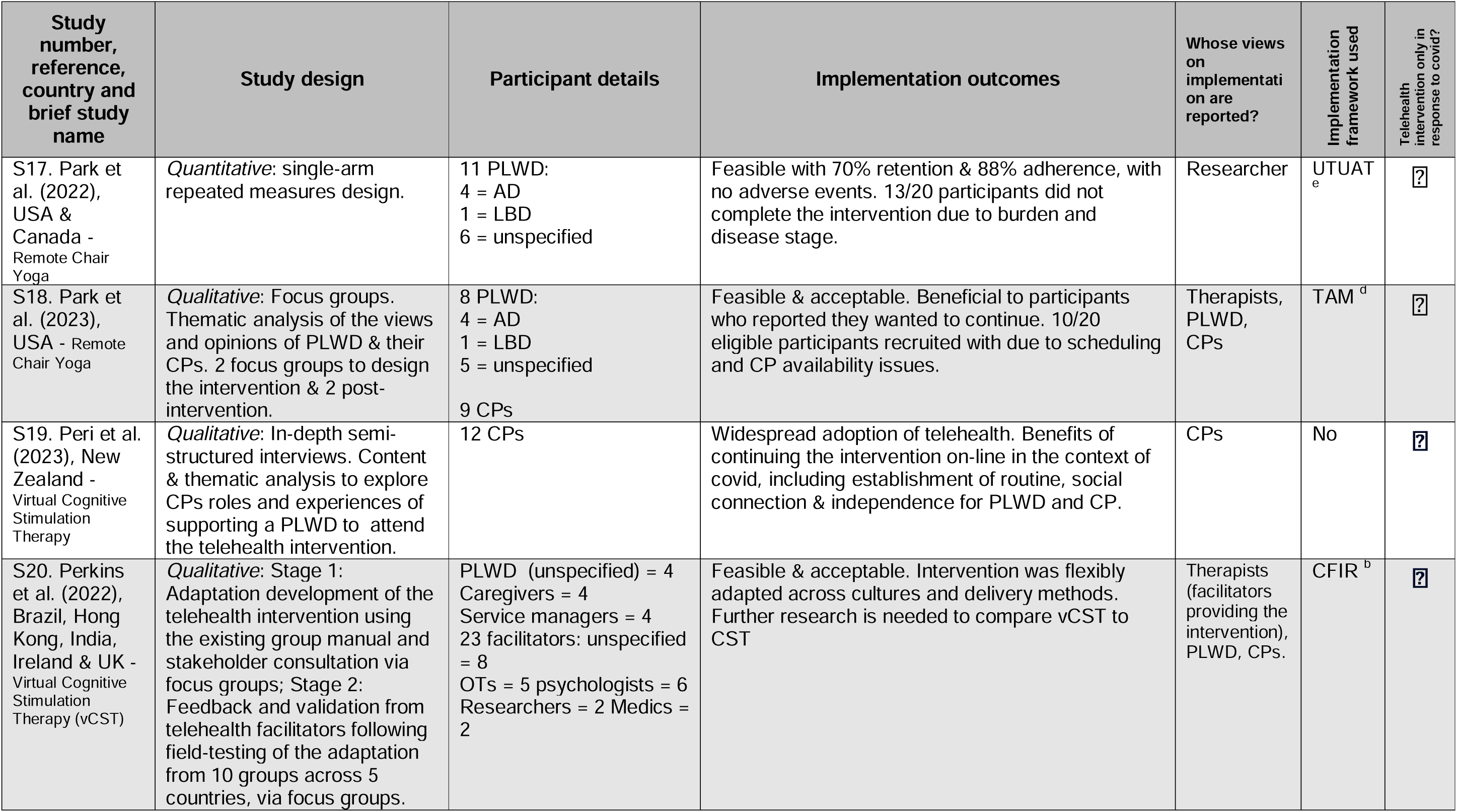
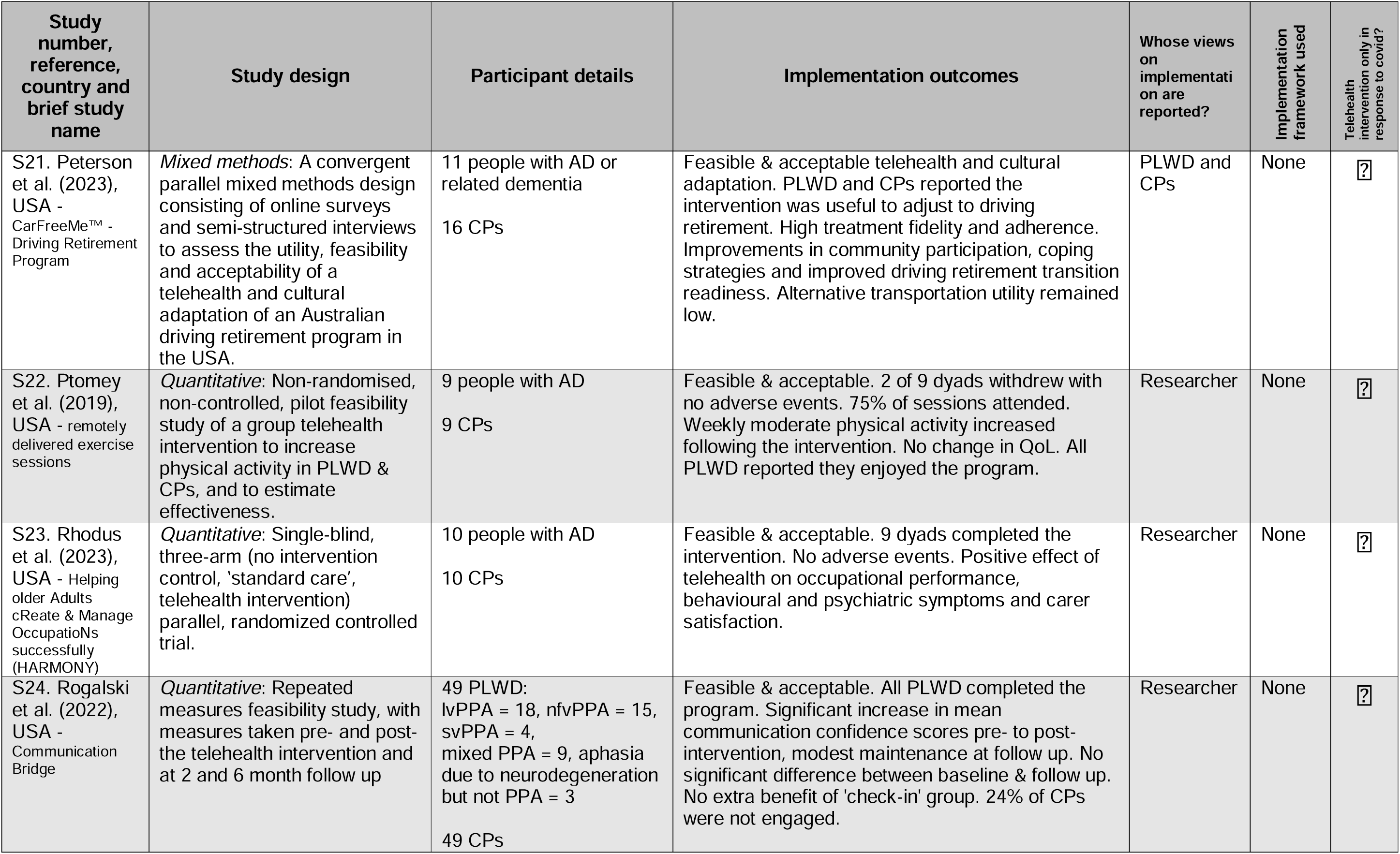
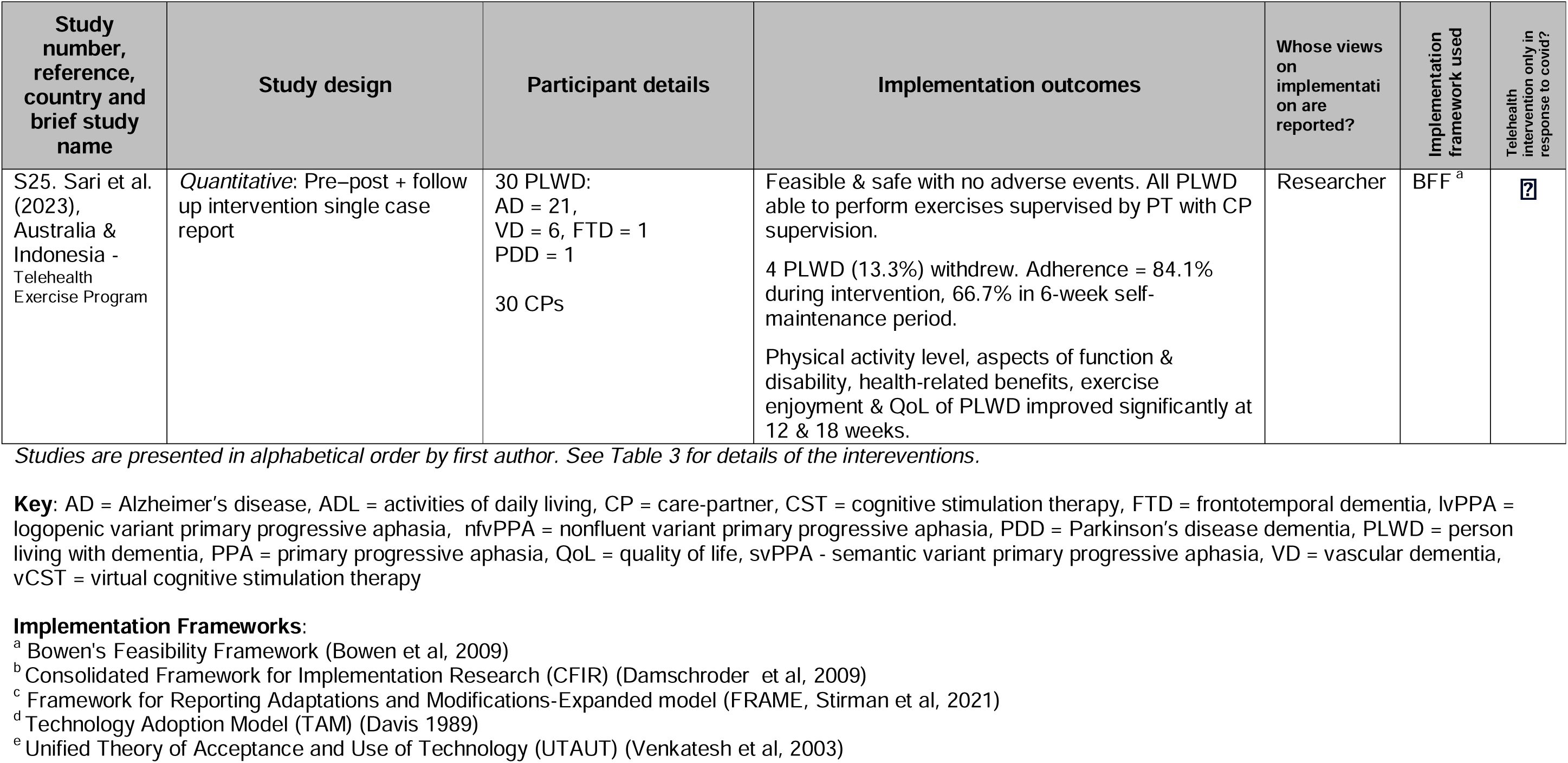
Study characteristics.

Six studies used frameworks to inform implementation, or help describe feasibility, adaptation and acceptance. Perkins et al. (2020) used the CFIR (Damschroder et al., 2009). Fredriksen-Goldsen et al. (2023) used the Framework for Reporting Adaptations and Modifications-Expanded model (Stirman et al., 2019); Park et al. (2022) used the Unified Theory of Acceptance and Use of Technology (Venkatesh et al., 2003); Park et al. (2021) and Lai et al. (2022) both used the Technology Adoption Model (Davis et al., 1989); and Sari et al. (2023) used Bowen’s Feasibility Framework (Bowen et al., 2009).

All 25 studies reported positive implementation outcomes. One study (Lai et al., 2022) reported possible negative family outcomes due to increased care partner support burden during the telehealth intervention. Views on implementation were reported based on the opinions of researchers (included in 14 studies), people living with dementia (in seven studies), care partners (nine studies) and therapists (eight studies).

No papers provided health economics data, or details of implementation costs. Despite this, seven studies (28%) made assumptions about cost savings of telehealth over in-person intervention delivery. These included time and money savings related to cancellations and session scheduling (Fredriksen-Goldsen et al., 2023), travel (Beeke et al., 2021; Menengiç et al., 2021), perceived efficiency (Di Lorito et al., 2021; Ptomey et al., 2019) and emotional cost savings such as reduced participant stress (Lai et al., 2022; Perkins et al., 2022).

The inclusion criteria in 17 studies (68%) resulted in digital exclusion (e.g., participants had to have their own equipment, or internet connection, or prior technological skill). Of the eight papers without this inclusion criteria, three studies provided equipment or training (Fanning et al., 2023, Fredriksen-Goldsen et al, 2023, Laver et al., 2022), with five studies providing no documented solution to support technology use (Henry et al., 2018, 2019, Jung et al., 2023, Peterson et al., 2023, Rogalski et al., 2022). Three (12%) studies excluded participants with a communication difficulty (Fay et al., 2023; Lai et al., 2022; Sari et al., 2023).

Ten studies (40%) described intereventions which were necessitated by the COVID-19 pandemic, i.e. they were not designed as telehealth studies from the outset, but were adapted for telehealth due to the pandemic (Beeke et al., 2021; Cowley at al., 2022; Di Lorito et al., 2021, 2022; Fanning et al., 2023); Fay et al., 2023; Fredriksen-Goldsen et al., 2023; Peri et al., 2023); Perkins et al., 2022; Sari et al., 2023).

### Intervention characteristics

The 25 retrieved studies described 20 different interventions. These are summarised in Table 3 according to the characteristics from the template for intervention description and replication (TIDieR) checklist (Hoffman et al., 2014). <u>Supplementary file 2</u> provides a complete summary of intervention components.

**Table 3:**
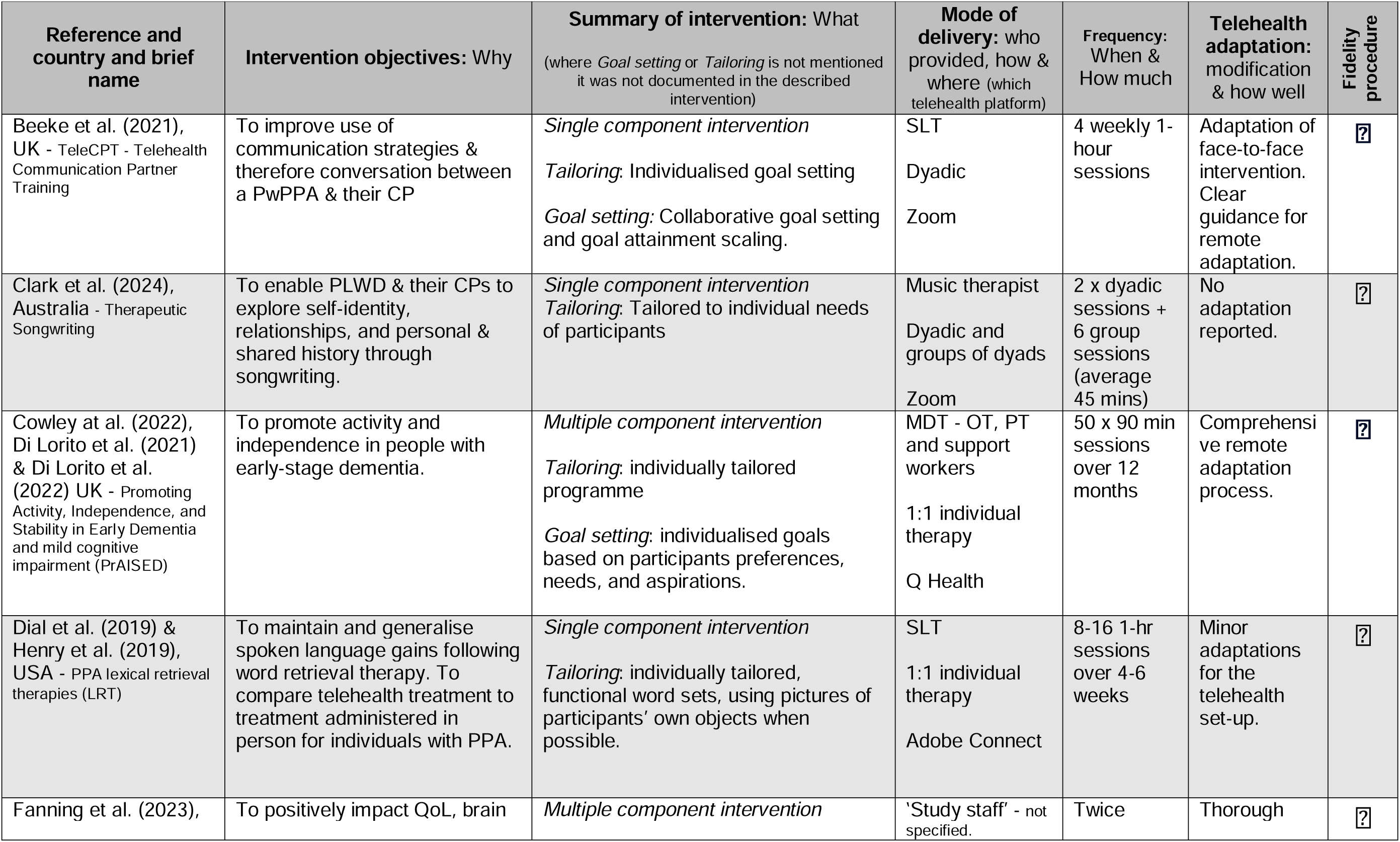
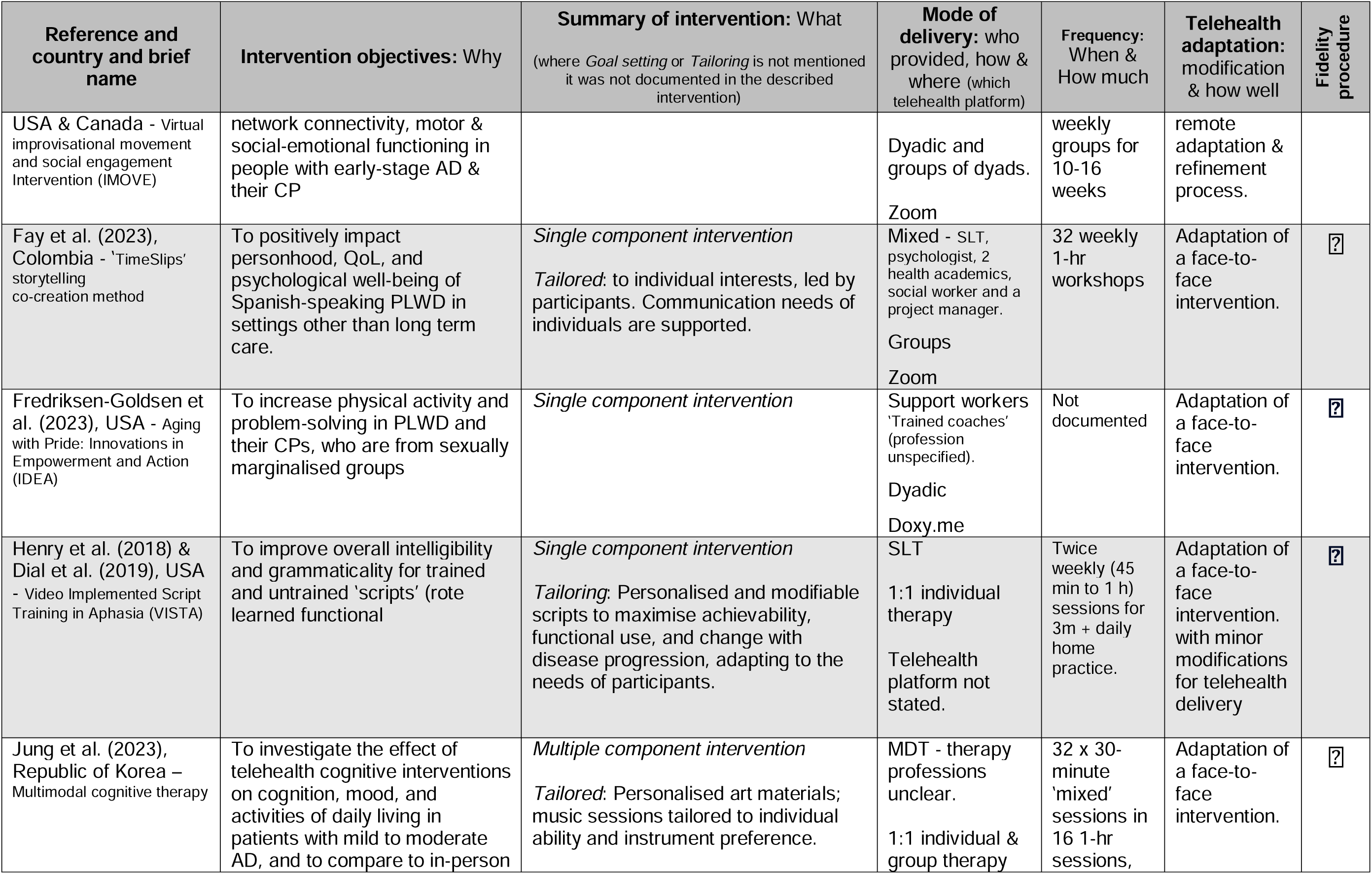
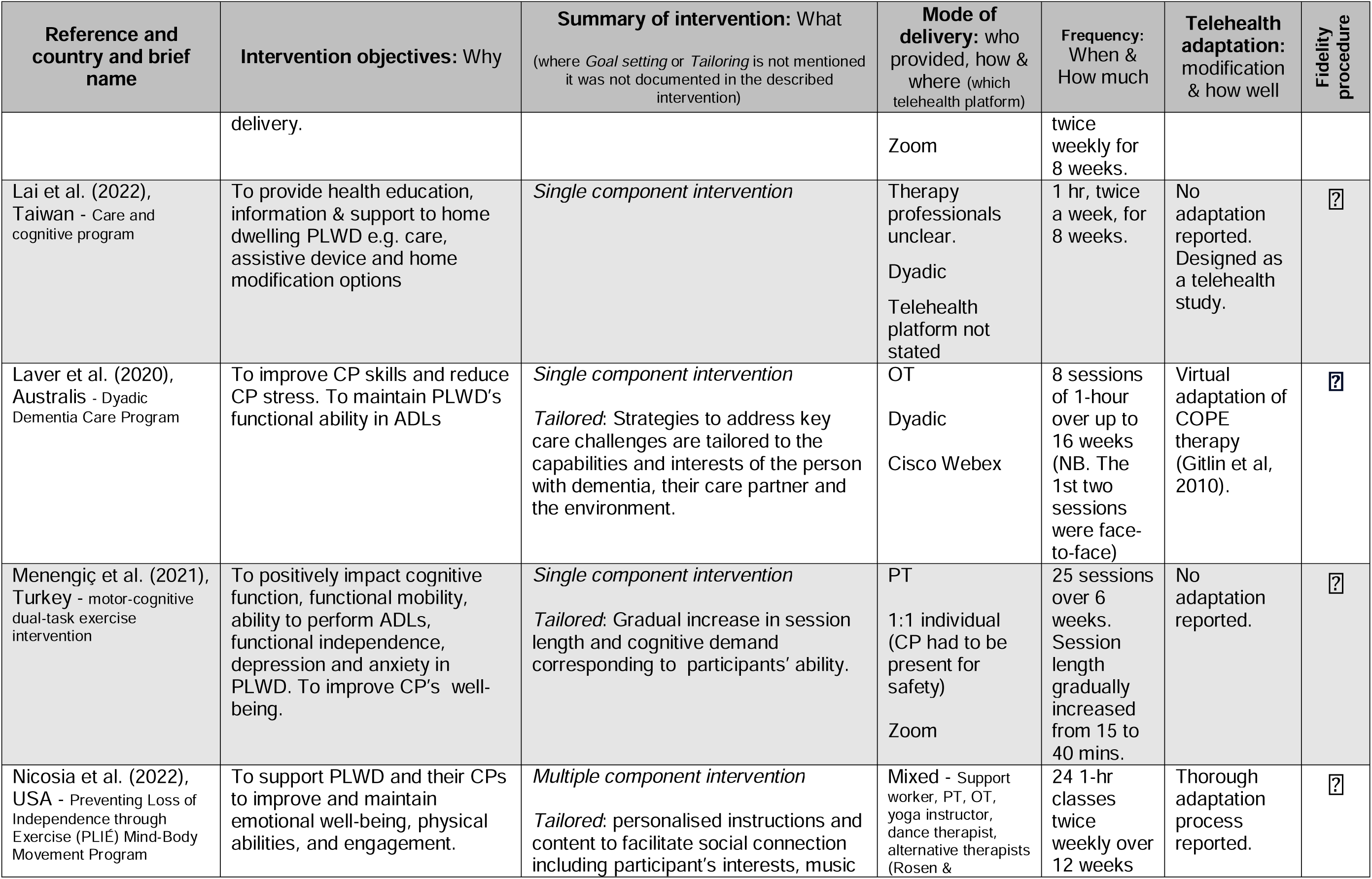
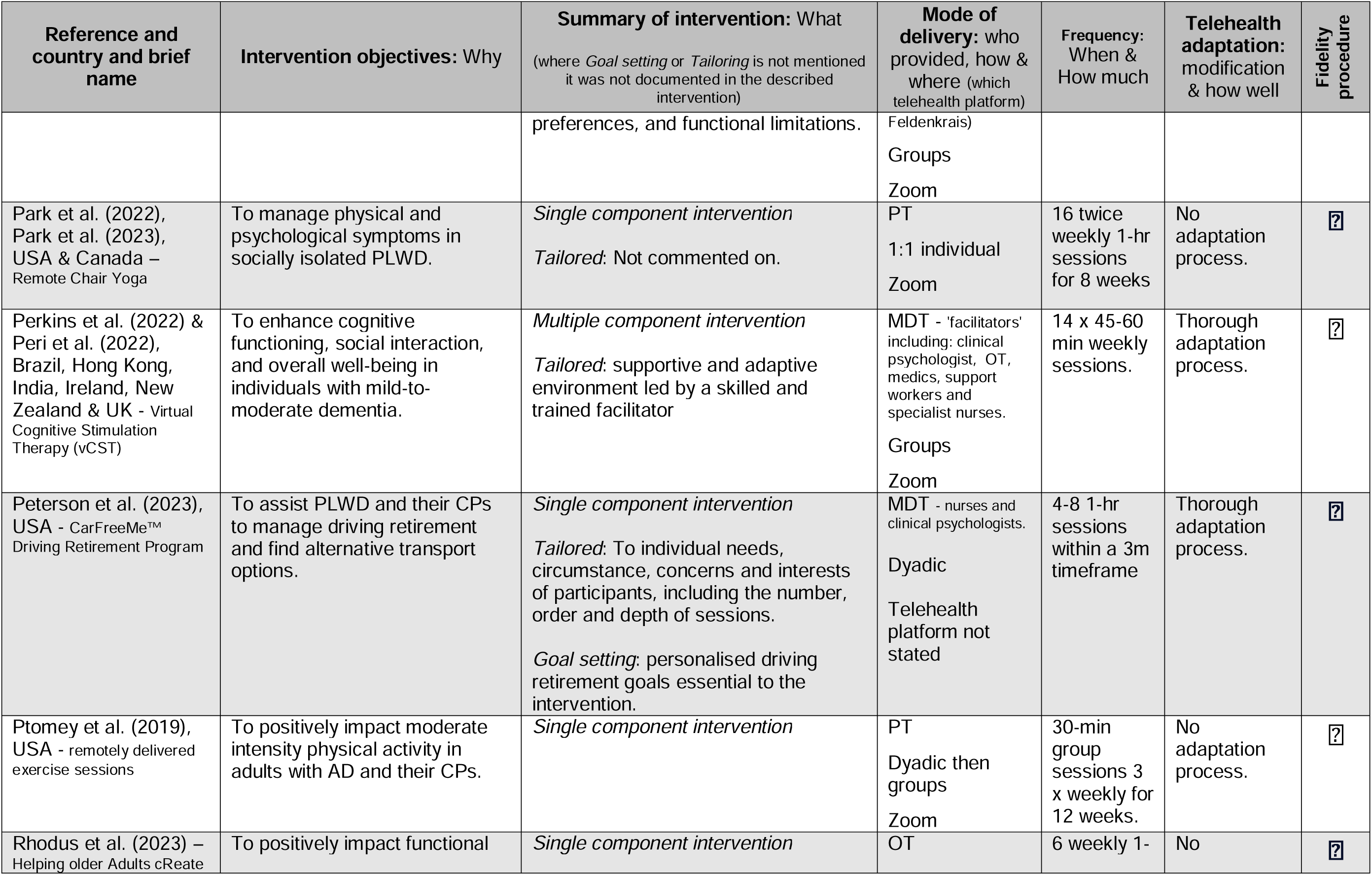
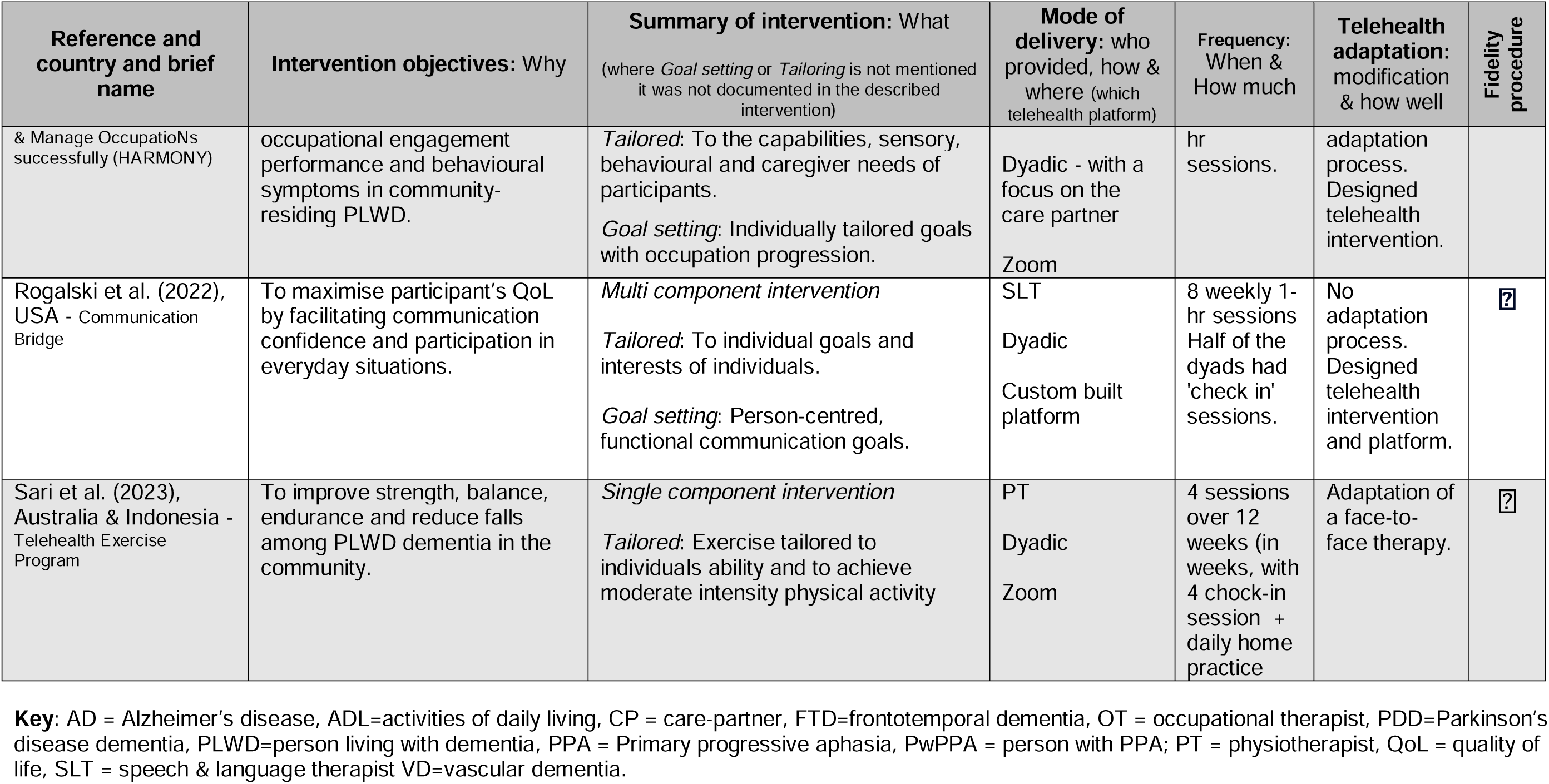
Intervention characteristics summary.

**Table 4:**
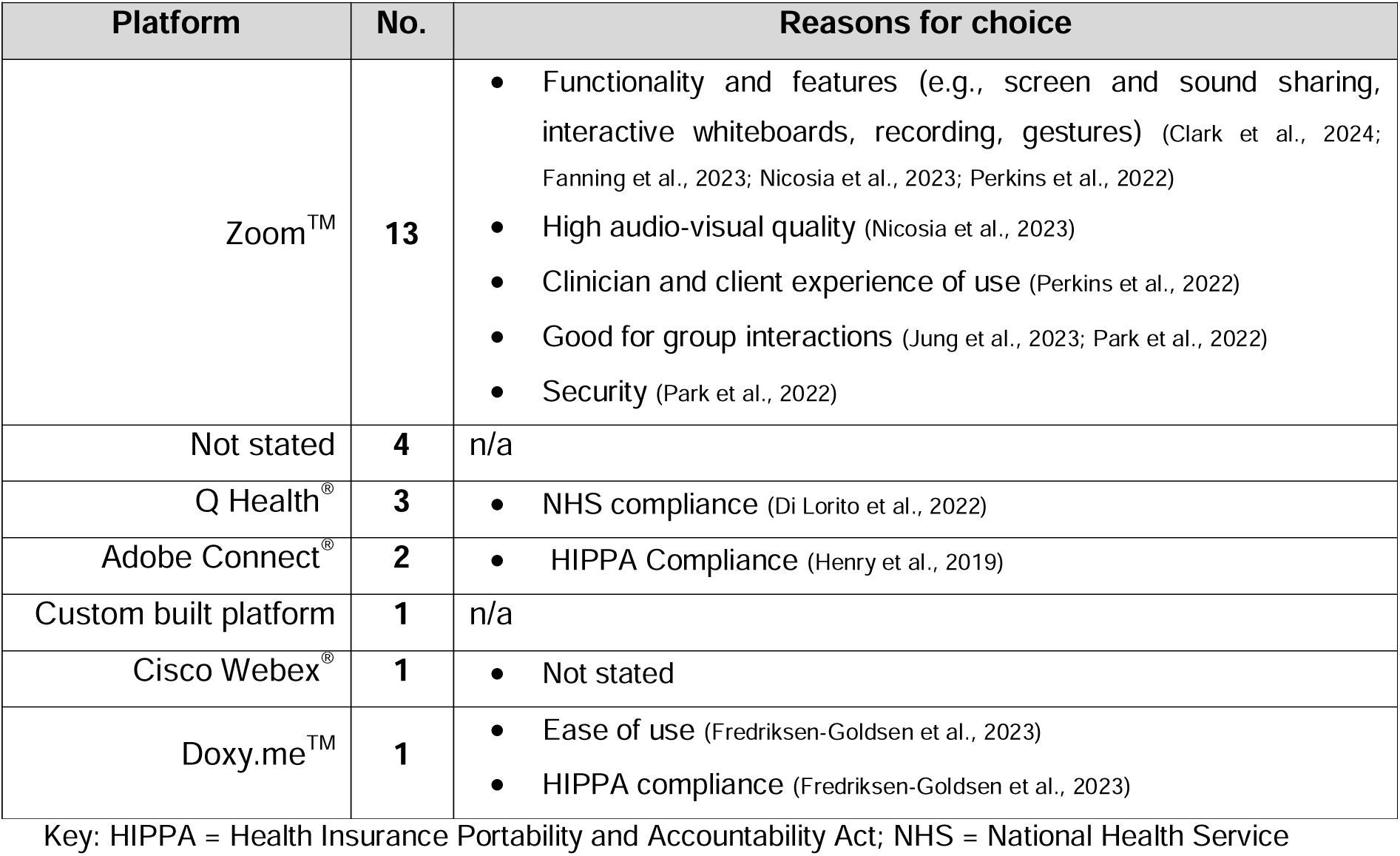
Videoconferencing platforms used.

| Platform | No. | Reasons for choice |
| --- | --- | --- |
| Zoom™ | 13 | • Functionality and features (e.g., screen and sound sharing, interactive whiteboards, recording, gestures) (Clark et al., 2024; Fanning et al., 2023; Nicosia et al., 2023; Perkins et al., 2022)<br>• High audio-visual quality (Nicosia et al., 2023)<br>• Clinician and client experience of use (Perkins et al., 2022)<br>• Good for group interactions (Jung et al., 2023; Park et al., 2022)<br>• Security (Park et al., 2022) |
| Not stated | 4 | n/a |
| Q Health® | 3 | • NHS compliance (Di Lorito et al., 2022) |
| Adobe Connect® | 2 | • HIPPA Compliance (Henry et al., 2019) |
| Custom built platform | 1 | n/a |
| Cisco Webex® | 1 | • Not stated |
| Doxy.me™ | 1 | • Ease of use (Fredriksen-Goldsen et al., 2023)<br>• HIPPA compliance (Fredriksen-Goldsen et al., 2023) |

Key: HIPPA = Health Insurance Portability and Accountability Act; NHS = National Health Service

#### Why (intervention objectives), what (intervention type) and who (intervention provider)

The 20 interventions represented a range of intervention types and providers. Six studies investigated multiple component interventions (e.g., physical therapy combined with cognitive therapy), five of which were delivered by MDTs (Cowley at al., 2022; Di Lorito et al., 2021, 2022, Jung et al., 2023; Nicosia et al, 2022; Perkins et al., 2022; Peri et al., 2022). The remaining multi component intervention was a complex intervention delivered by speech and language therapists, which included disease education and dyad support components in addition to a variety of communication therapy approaches (Rogalski et al., 2022). The remaining fourteen studies investigated single component interventions (e.g., communication, or physical therapy focused alone).

In summary, five interventions were predominantly communication therapies delivered by speech & language therapists (Beeke et al., 2021, Dial et al., 2019., Henry et al., 2018, 2019, Rogalski et al., 2022), five were physical therapies delivered by physiotherapists (Menengiç et al., 2021, Park et al, 2022, 2023, Ptomey et al., 2019, Sari et al., 2023), two focussed care and environment optimisation delivered by occupational therapists (Laver et al., 2020, Rhodus et al., 2023), and one described a collaborative songwriting intervention delivered by music therapists (Clark et al., 2024). Five interventions were delivered by MDTs, which included allied health professionals, psychologists and complementary therapists (e.g., yoga and Feldenkrais instructors). Of these, two were cognitive rehabilitation therapies (Jung et al., 2023; Perkins et al., 2022; Peri et al. 2022), and three were a mixture of cognitive, physical, complimentary and communication therapies (Cowley at al., 2022; Di Lorito et al., 2021, 2022; Nicosia et al., 2022). In the remaining three interventions it was unclear who delivered the intervention. These were a cognitive behavioural intervention (Fredriksen-Goldsen et al., 2023), a health education intervention (Lai et al., 2022) and an improvisational movement and social engagement intervention. (Fanning et al., 2023). Of all 20 reported interventions a quarter (n=5) documented personalised goal setting processes (Beeke et al. 2021; Cowley at al., 2022; Di Lorito et al., 2021, 2022; Peterson et al., 2021; Rhodus et al., 2023; Rogalski et al., 2022)

#### How (delivery mode and configuration) and where (telehealth platform)

Six interventions (30%) were one-to-one between a therapist and a person with dementia (Cowley at al., 2022, Dial et al., 2019, Di Lorito et al., 2021, 2022, Henry et al., 2018, 2019, Jung et al., 2023, Menengiç et al., 2021, Park et al., 2022). Nine and the majority of interventions (45%) were dyadic, i.e., the person with dementia and their care partner engaged together with a therapist (Beeke et al., 2021; Sari et al., 2023; Fredriksen-Goldsen et al., 2023; Lai et al.; 2022, Laver et al., 2022; Peterson et al., 2023; Ptomey et al., 2019, Rhodus et al., 2023; Rogalski et al., 2022; Sari et al., 2023). The remaining five interventions (25%) were group interventions (Fay et al., 2023, Nicosia et al., 2022, Perkins et al., 2022, Peri et al., 2022) including two groups of dyads (Clarke et al., 2024, Fanning et al. 2023).

Most telehealth interventions used the videoconferencing platform Zoom^TM^ (Zoom Video Communications Inc., 2024) (n=13) (Beeke et al., 2021, Clark et al., 2024, Fanning et al., 2023, Fay et al, 2023, Jung et al., 2023, Menengiç et al., 2021, Nicosia et al., 2022, Park et al., 2022, 2023, Perkins et al., 2022, Peri et al., 2022, Rhodus et al., 2023, Sari et al, 2023). Four interventions did not specify which videoconferencing platform they used (Dial et al., 2019, Henry et al., 2018, Lai et al., 2022, Peterson et al., 2023). Only one intervention used a custom built web application (Rogalski et al., 2022), the rest used publicly available technologies. See Table 5 for a summary of telehealth platforms used and why.

**Table 5:**
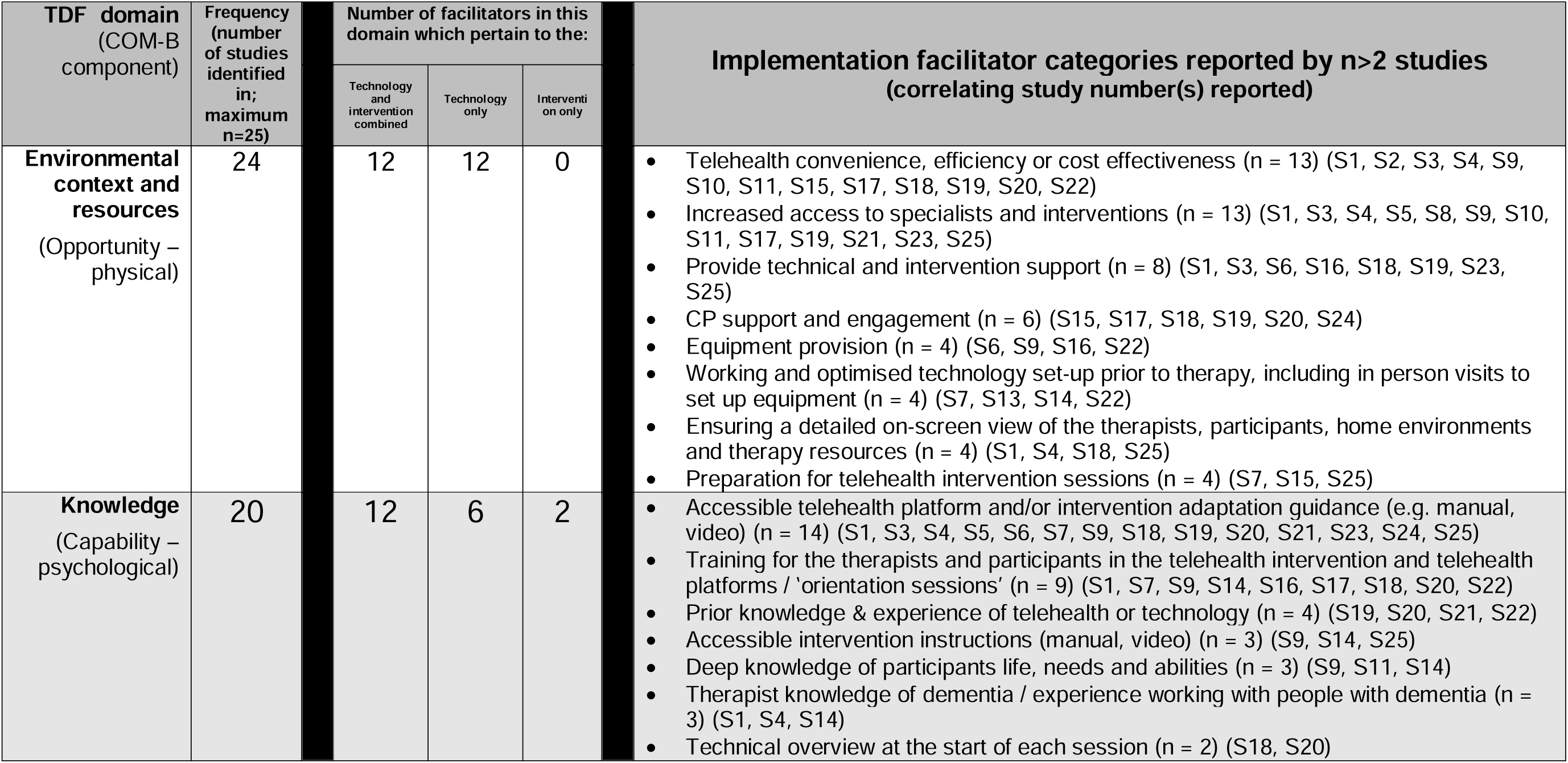
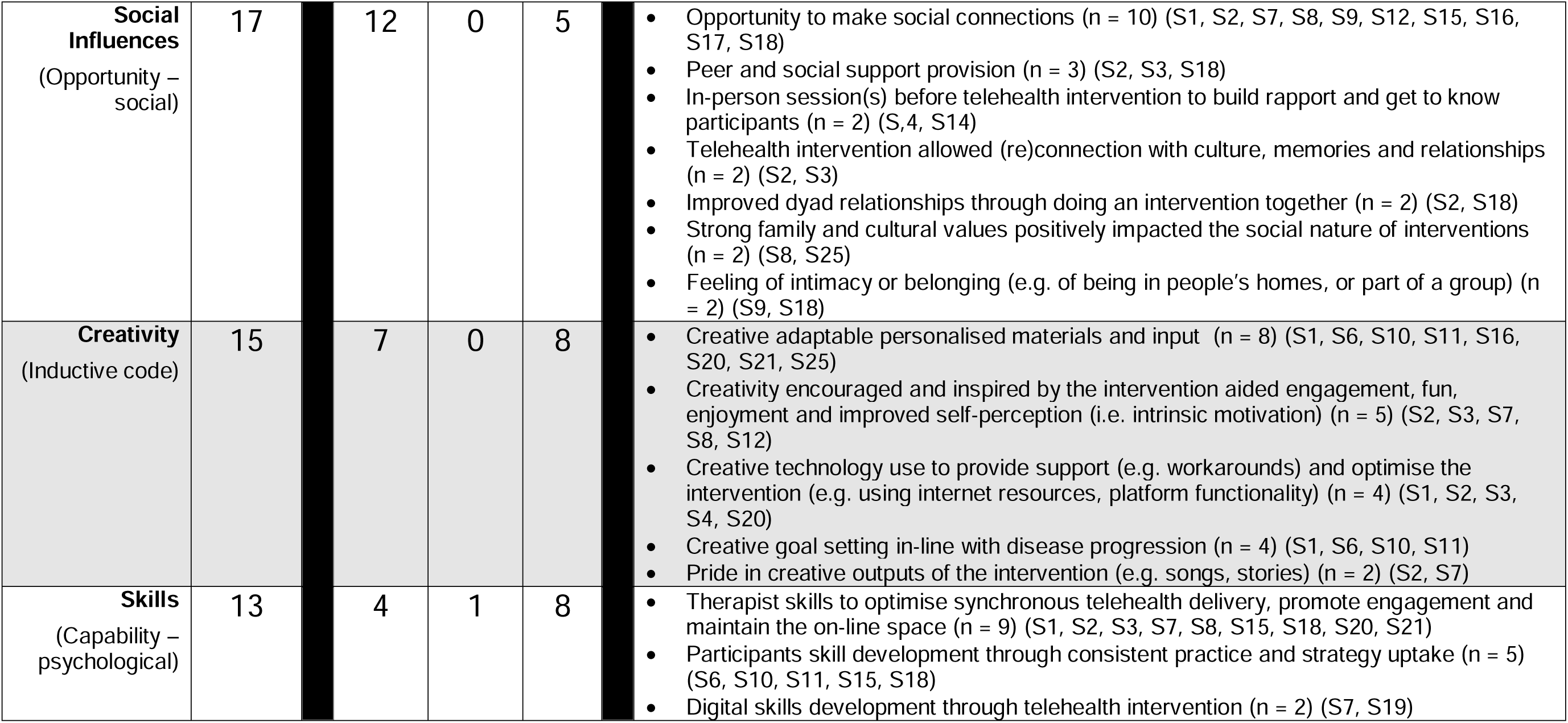
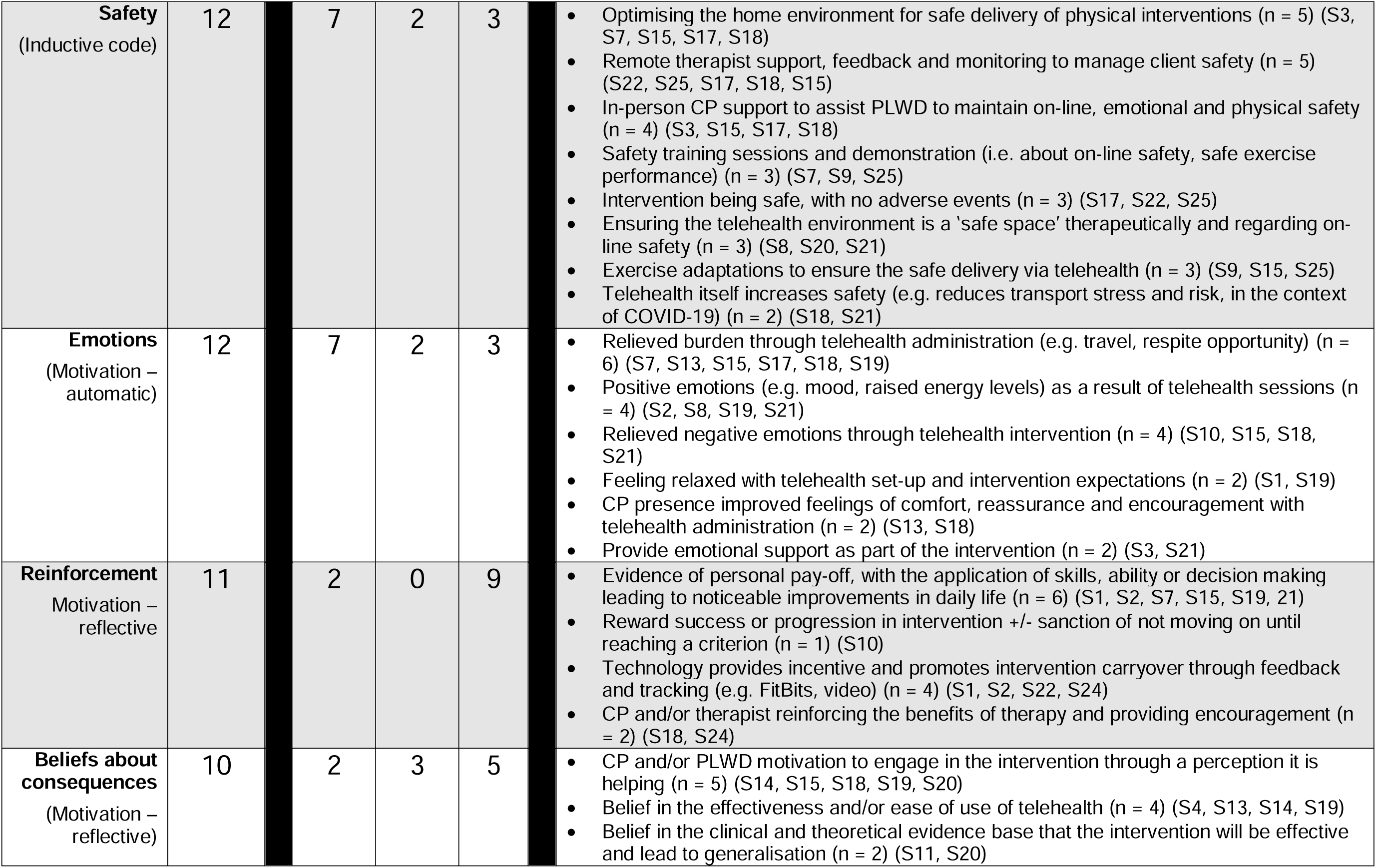
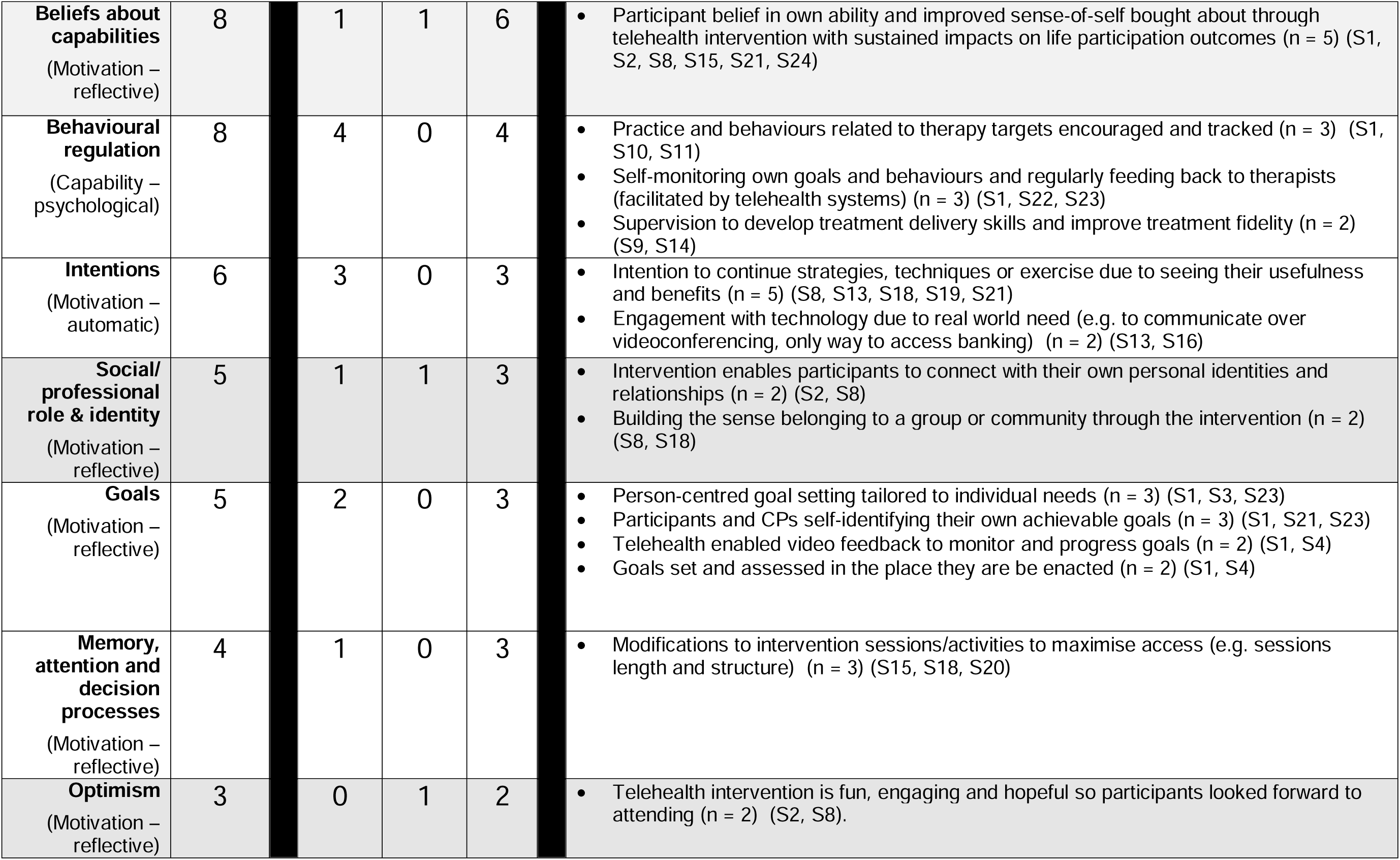
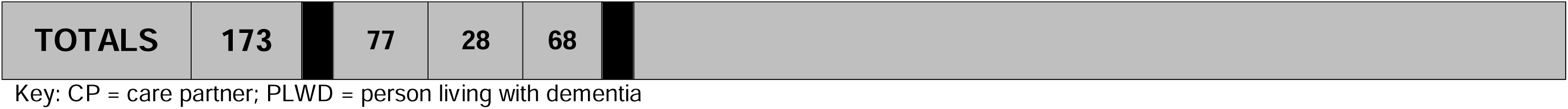
Telehealth intervention implementation facilitators identified within the TDF, COM-B and inductively derived domains, with a summary of identified facilitators.

#### When and how much (duration, number of sessions)

There was a wide range of intervention durations and intensities reported, from four one-hour sessions delivered once-weekly, (Beeke et al., 2021) to 50 ninety-minute sessions over twelve months (Di Lorito et al., 2021).

#### Modifications and how well (adaptations and fidelity)

Seven interventions (35%) were designed and trialled as telehealth interventions from the outset. The remaining thirteen interventions described telehealth adaptations of a face-to-face intervention (65%). Eight interventions (40%) reported fidelity procedures.

### Implementation barriers and facilitators

#### TDF domains

Implementation determinants were deductively categorised into the 14 TDF domains (see Tables 5 and 6 for a comprehensive list of TDF domains). Two further domains ‘Creativity’ and ‘Safety’ were inductively identified in the content analysis. These domains included determinants related to:

- *Creativity*: Creative telehealth adaptation such as therapist creativity to overcome technological or contextual barriers in the remote delivery of an intervention, creative use of technology to enhance an intervention, creative goal setting and personalised materials in-line with individual needs and disease progression, and pride from creative output of interventions.
- *Safety*: Participant physical, psychological or digital safety to engage in a telehealth intervention without a therapist present in-person; feelings of a ‘safe space’ engendered by a group or therapeutic alliance.

**Table 6:**
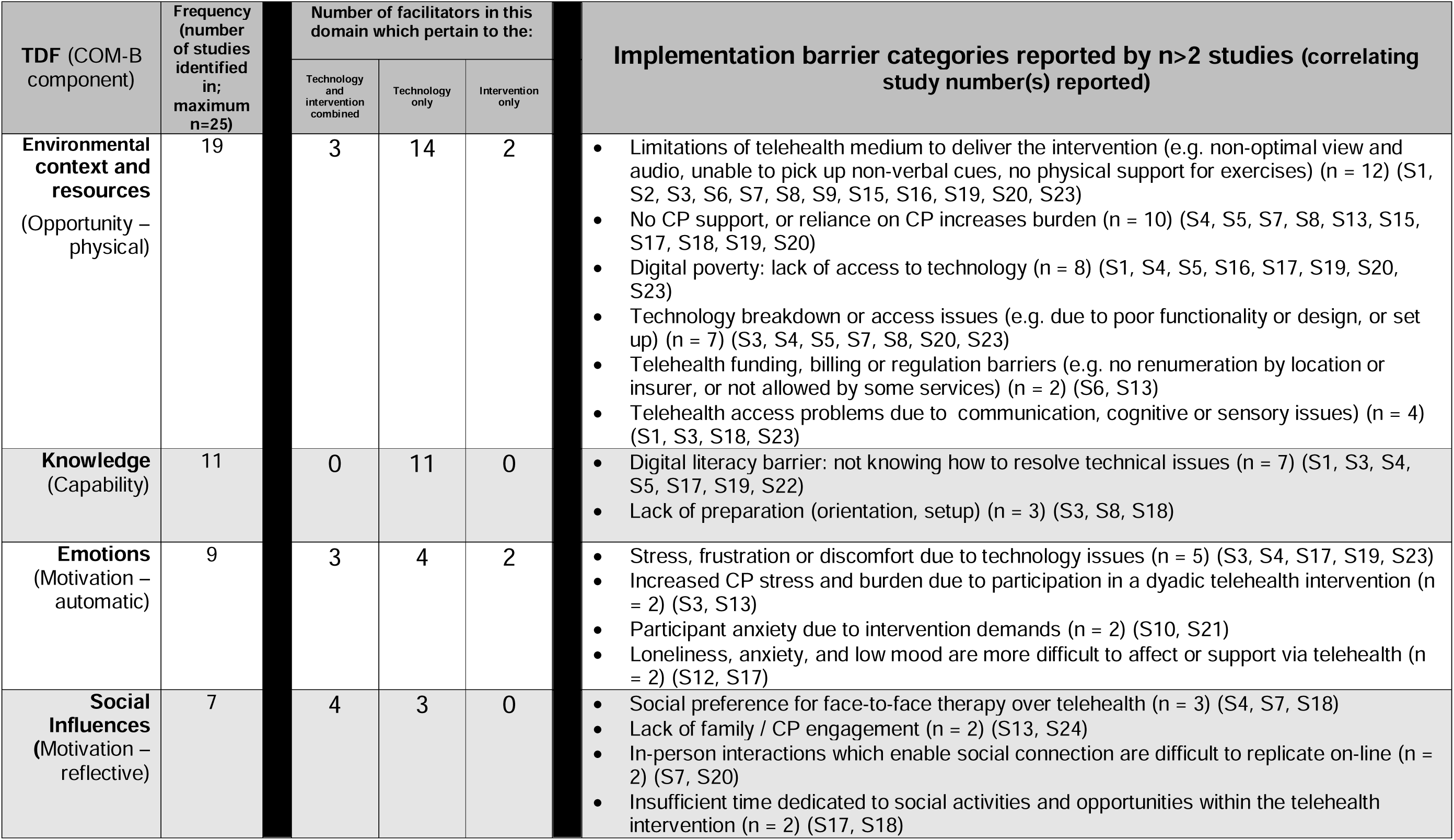
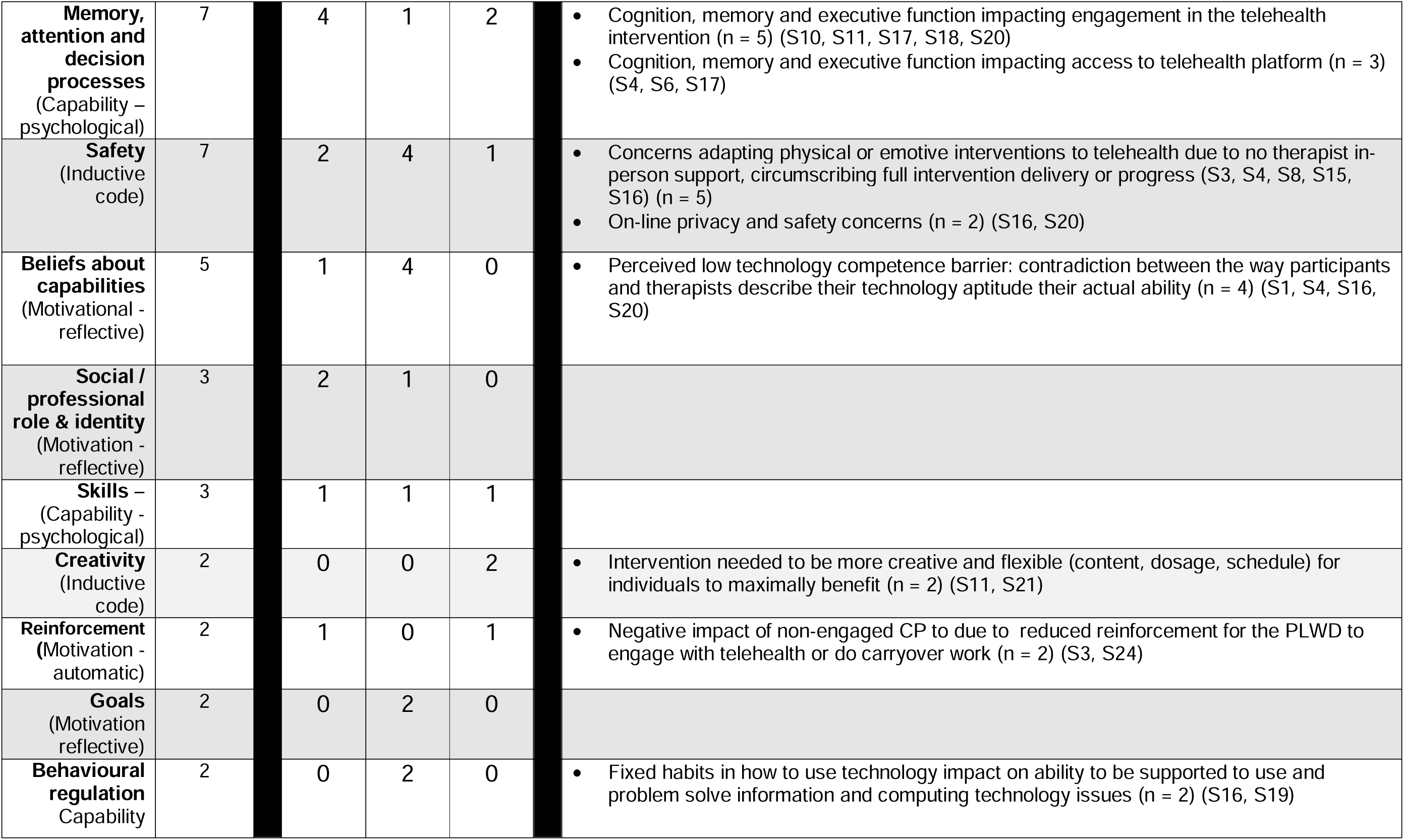
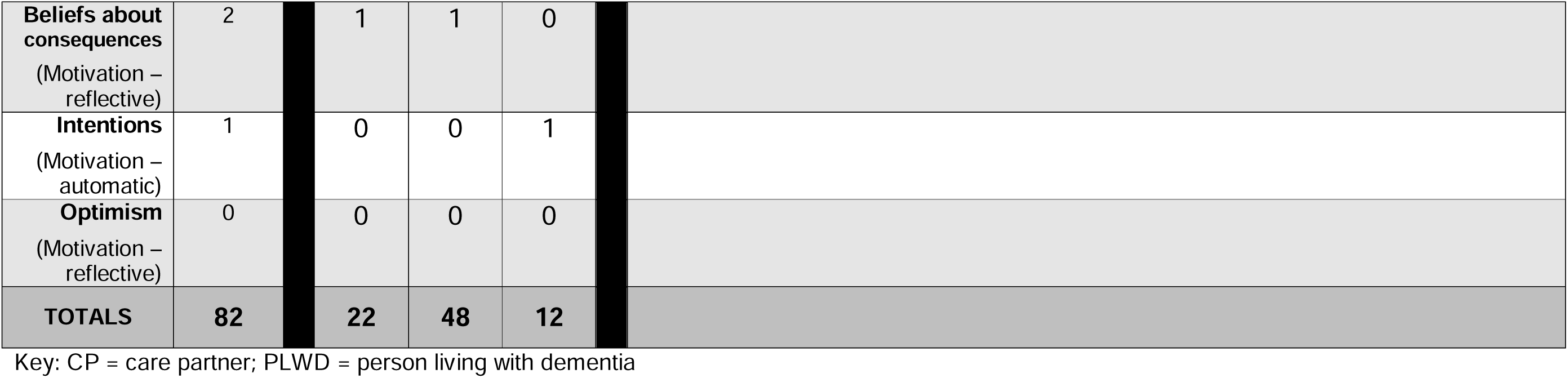
Telehealth intervention implementation barriers identified within the TDF, COM-B and inductively derived domains, with a summary of identified barriers.

Frequently identified determinants relating to *creativity* and *safety* did not fit neatly within TDF domains, warranting the creating two new inductive domains to capture these nuances.

In total, across 25 studies, facilitator domains were identified 173 times. Of these, 68 related to the intervention itself, 77 pertained to an aspect of telehealth implementation (i.e., technology and intervention combined) and 28 pertained to the technology only. Facilitator domains are summarised in Table 5 alongside exemplar categories which appear in more than one paper (n ≥ 2). Barrier domains were identified 83 times. Of these, 48 pertained to the technology, 22 to telehealth implementation and 12 pertained only to the intervention. Barrier domains are summarised in Table 6 alongside exemplar categories which appear in more than one paper (n ≥ 2). For full detail of extracted determinants from each paper, synthesised into the 14 TDF plus two 2 additional domains see the <u>supplementary file 3.</u>

### COM-B concepts

Subsuming TDF domains within the COM-B model (see Table 7 below) shows that *Motivation* component determinants are the most frequently identified facilitators (n = 60). Specific examples include participant motivation from seeing real life intervention benefits through feedback and reflection, or therapist beliefs in the effectiveness of telehealth. *Opportunity* component determinants are the most frequent barriers (n = 26). Specific examples include access issues due to digital poverty and limitations of the telehealth context that prevented full protocol delivery. *Capability* barriers most frequently related to knowledge or training needs on how to use technology once in place.

**Table 8:** Summary of telehealth intervention implementation determinants within COM-B components.

|  | COM-B domain |  | Overlap with TDF domains | Total |  |
| --- | --- | --- | --- | --- | --- |
| Facilitators | Capability | Psychological | 44 | 44 | 146 |
|  |  | Physical | 0 |  |  |
|  | Opportunity | Social | 17 | 41 |  |
|  |  | Physical | 24 |  |  |
|  | Motivation | Automatic | 19 | 60 |  |
|  |  | Reflective | 42 |  |  |
| Barriers | Capability | Psychological | 23 | 23 | 73 |
|  |  | Physical | 0 |  |  |
|  | Opportunity | Social | 8 | 26 |  |
|  |  | Physical | 18 |  |  |
|  | Motivation | Automatic | 12 | 24 |  |
|  |  | Reflective | 13 |  |  |

## Discussion

This systematic review identified 25 peer reviewed studies which investigated barriers and facilitators to implementing synchronous telehealth interventions with people with dementia. Despite categorising determinants using the TDF and COM-B theoretical frameworks, two additional domains ‘Safety’ and ‘Creativity’ were identified. The data were further categorised as either barriers or facilitators to implementation and classified based on whether they pertained solely to the intervention, the technology, or both.

Studies in this review did not consistently report on all aspects of telehealth interventions and their implementation. Only six of 25 (24%) studies used a framework to report on implementation or adaptation processes, which may have impacted on reporting quality and completeness. Four studies (16%) did not report which videoconferencing platform they used, two studies did not specify who delivered the intervention and 12 studies did not document dementia type. Only four studies documented equipment provision to overcome digital exclusion (Dial et al., 2019; Fredriksen-Goldsen et al., 2023; Nicosia et al., 2022; Ptomey et al., 2019). Ten of the 25 included studies (40%) reported on fidelity, a key implementation outcome (Proctor et al., 2011). Although this compares favourably to evidence of fidelity report ting in the aphasia (21%; Brogan et al., 2019) and dementia (22.5%; Hwang et al., 2024) behavioural intervention literature, this is further evidence of inconsistent reporting. Inconsistent reporting and inadequate specification have been shown to compromise the replicability and efficacy of complex interventions and their implementation (Hoffman et al., 2014; Lewis et al., 2018; Michie et al., 2009; Proctor et al., 2023; Stirman et al., 2019). Including details of telehealth platform used, how it was adapted to facilitate the intervention, costs, and who delivered and benefitted from the intervention, can facilitate future implementation efforts across contexts (Miller et al., 2021).

Care partner involvement and support were identified as important facilitators to telehealth intervention implementation, while their lack of such involvement and support acted as a corresponding barrier. Care partners were important in maintaining safety, helping overcome technological barriers and to providing motivational support. Care partner engagement was a particularly important implementation determinant linked to clinical and functional outcomes in Rogalski et al.’s (2022) multi-component SLT telehealth intervention. Care partner involvement has also been identified as an important implementation determinant in quality of life interventions in dementia (Hockley et al., 2023; Wyman et al., 2022). A crucial implementation strategy may therefore be to encourage and support care partner input. Dyadic interventions may be particularly amenable to telehealth interventions, and to those living with dementia, hence the high representation of dyadic interventions in this review.

Ease of use of the telehealth platform is an important determinant in implementation. Zoom was the most frequently used platform. This is perhaps unsurprising considering its ease of use compared to other videoconferencing software, and widespread usage in telehealth (Hersh, 2024; Hilari et al., 2024). Common determinants identified in this review highlighted the importance of optimal human-computer interaction (HCI) design and access, replicating findings from a previous telehealth review (Scott Kruse et al., 2016). HCI design is the process of creating user-friendly digital systems that are efficient and accessible by focusing on how people interact with the technology. Unfortunately, platforms used by health care organisations can have poor HCI design and access (e.g. Di Lorito et al., 2021). Therefore, a key implementation strategy is engagement with information governance managers in healthcare systems for use of preferred platforms such as Zoom (e.g. Peri et al, 2023).

The importance of replicating social aspects of face-to-face interventions via telehealth was reflected by the prominence of the *Social influences* domain as a facilitator, the third most frequently identified facilitator domain. Commonly identified determinants included incorporating social opportunities into intervention schedules (e.g., Park et al., 2023), incorporating local cultural elements into interventions (e.g., Fay et al., 2023), and meeting face-to-face before embarking on remote therapy (Di Lorito et al, 2021; Laver et al, 2020). The importance of peer and social support is well documented for people with dementia and their care partners (Carter et al., 2020; Willis et al., 2016), including providing such support via telehealth (Dam et al., 2016), including those with rare dementias (Harding et al., Sullivan et al., 2022). As a caution against overinterpretation, the importance of social factors may be inflated in studies conducted during the COVID-19 pandemic, when telehealth interventions themselves may have provided rare social opportunities for participants. Nevertheless, the importance of digital platforms for people living with dementia to counter the stresses of the pandemic through facilitating social connection and self-actualisation has been previously reported (Talbot et al., 2022). Social facilitators have also been reported outside the pandemic context, for example in telehealth peer-support groups for care partners of individuals with primary progressive aphasia (Schaffer & Henry, 2023), and in previous systematic reviews of telehealth in dementia (Liang & Aranda, 2023; Ye et al., 2025). Ensuring that social and peer support opportunities prevalent in in-person interventions are not lost in telehealth delivery may be important to implementation and sustainability.

*Creativity* was repeatedly identified (n=15 studies) in this review as a facilitator domain. *Creativity* was often related to the *Skills* (the 4^th^ most frequently identified domain) of therapists to adapt materials and interventions for on-line use, make the most of opportunities provided by technology and overcome telehealth access barriers. The Royal College of Speech & Language Therapists surveyed members on changes to service delivery made during the COVID-19 pandemic and found adoption of telehealth was one of the most common, with over half of speech and language therapists reporting this provided opportunities for innovative, creative practice (61%) and skills development (54.4%) (RCSLT, 2020). *Creativity* also encompassed determinants related to the adaptability of interventions for telehealth delivery. Given that ten studies employed telehealth only in response to the COVID-19 pandemic, the prominence of *Creativity* may be overrepresented in studies from this period. Whenever *Creativity* was identified as a facilitator, so was the *Goal domain,* implying a connection. However, patient centred goal setting was only documented in five interventions. This is likely related to the strict therapy protocols seen in several interventions, which required repetitive practice but with the absence of personally relevant goals (e.g., Ptomey et al., 2023; Sari et al., 2023). A consideration for future implementation studies of telehealth dementia interventions should be the inclusion and adaptability of goal-setting procedures, so that goals can be modified in line with disease progression and remain relevant and useful to people’s changing lives, thus improving motivation.

Most barriers identified related to technology and sit within *Opportunity* and *Capability* COM-B components. Utilising the Cynefin framework, these determinants could often be classified as *clear* benefitting from relatively straightforward implementation strategies to address them (e.g., providing equipment, providing training and support on how to use that equipment). It may therefore be sensible to address these barriers first, before testing the feasibility of a telehealth intervention, to better ensure implementation outcomes are not impacting intervention outcomes. Notably, considering four of the included telehealth interventions were implemented in what can be considered the global south, no papers addressed possible contextual determinants of underdeveloped digital infrastructure. This may be related to the high proportion of studies which already had inclusion criterea which resulted in digital exclusion, i.e., people in digital poverty could not access these therapies, or they were not implemented in under resourced areas. Broadening inclusion to telehealth therapy opportunities, especially in ‘need to reach’ areas, would give a more rounded picture of telehealth implementation determinants. Broadening inclusion and augmenting sample diversity have been recommended in previous dementia telehealth in dementia reviews (Liang & Aranda, 2023; Ye et al., 2025).

Conversely, telehealth intervention facilitators were mostly to do with the intervention itself and mainly sit within the *Motivation* COM-B component. These could be conceptualised as *complex* determinants. For example, therapy engagement can be promoted by dynamically and creatively modifying therapeutic input to align with changing individual goals, circumstances, and relationships in line with disease progression (e.g., Cowley et al., 2023, Rhodus et al., 2023). This aligns with the main themes of person-centredness and complexity, highlighted by the international expert consensus of the principles and philosophies for working with people with people with PPA (Volkmer et al., 2023b). It also aligns with the MRC guidance on developing complex interventions, which views interventions as dynamic and evolving within settings (Skivington et al., 2021). Complex implementation strategies, such as investigating motivation, should therefore be a priority for further investigation during feasibility testing.

Furthermore, if ‘clear’ implementation strategies have already been put in place it may impact ‘complex’ determinants. For example, *Knowledge* and *Resource* domains may interact with the *Social/professional role and identity* domain. *Social/professional role and identity* contains determinants including therapists professional identity being challenged through not looking competent using technology, a determinant also identified in previous telehealth implementation literature (Oudbier et al., 2024). Prioritising technology provision (*Resource*) and training (*Knowledge*) may then impact downstream motivational determinants.

The views of therapists themselves are underrepresented in the studies in this review. For example, eight of 25 studies included therapists’ views; however, even fewer studies identified therapists’ views within the domains of *Social/professional role and identity* (n=2) or therapists’ *Beliefs about [the] consequences* of telehealth interventions (n=5). Although *‘Motivation’* component domains were the most frequently identified TDF domains overall, their significance in the implementation of telehealth interventions for dementia may still be underestimated. Future research should therefore probe individual *motivation* TDF domains more deeply, particularly considering their potential interaction with frequently identified TDF domains, which may be crucial to implementation and intervention success.

Many of the determinants identified in this review may not be unique to dementia interventions and appear broadly applicable across telehealth contexts and with other conditions. This highlights a paucity of determinants in the *Memory, Attention, and Decision-Making* domain, where more dementia-specific barriers might reasonably be expected. Finally, the limited use of implementation frameworks in designing and describing this set of telehealth interventions may contribute to a blurred boundary between determinants, potential implementation strategies and actual intervention components themselves. Greater use of implementation frameworks, and hybrid implementation–intervention designs, may help to better delineate this fuzzy boundary (Curran et al., 2012, 2022, Landes et al., 2019).

## Strengths & limitations

*Environmental context and resource* and *Knowledge* domains are commonly related to technology provision and use and have consistently been identified as determinants of telehealth intervention implementation (Borges do Nascimento et al., 2023; Oudbier et al., 2024). These domains are also predominately identified in TDF informed systematic reviews of interventions in healthcare (e.g., Mather et al., 2022; Schrubsole et al., 2021). Whilst it is possible that there is bias in the domains that the TDF foregrounds when used as an analysis tool, it also provides confidence in the current study’s methodology. This study made provision for this by using the COM-B model in conjunction with the TDF.

No papers were excluded from the current study based language. In reality, the search terms were in English, rendering it unlikely any non-English appears would be found. Indeed, only one paper not written in English was found (Yang et al., 2022) but the paper was screened out at the abstract/title stage as it was a meta-analysis. To improve equality, diversity and inclusion of studies in future systematic reviews, including search terms in languages other than English should be considered.

Only one researcher (RT) completed the quality assessment and inductive data coding. This may be considered a limitation. However, screening and data extraction was otherwise rigorously undertaken my multiple researchers providing strength and rigour.

## Conclusion

This study highlights the importance of systematic reporting of telehealth interventions in dementia to reduce barriers to implementation. The use of theory enabled understanding of implementation determinants and how they may interact, and of potential implementation strategy prioritisation. Most implementation barriers in telehealth dementia interventions were to do with technology. These barriers were often clear problems, relating to *Capability* and *Opportunity*. Most implementation facilitators were to do with the telehealth interventions themselves, relating to *Motivation*. Leveraging these facilitators is complex, requiring complex implementation strategies. Understanding how to manage complexity and unpredictability is a key implementation goal when providing telehealth interventions for people with dementia.

## Funding

AV and RT are funded by AV’s National Institute for Health Research (NIHR) Advanced Fellowship (NIHR302240)

CH is supported by the Alzheimer’s Society (AS-PhD-19a-006)

RV is supported by a Leverhulme Research Fellowship (RF-2023-690\10)

JDW has received funding support from the Alzheimer’s Society, Alzheimer’s Research UK, the Royal National Institute for Deaf People (Discovery Grant G105_WARREN), the NIHR University College London Hospitals Biomedical Research Centre, and The National Brain Appeal (Frontotemporal Dementia Research Studentship in Memory of David Blechner)

## Disclosure statement

The authors report there are no competing interests to declare.

## Data availability statement

The data that support the findings of this study are openly available in the open science framework repository at https://osf.io/gkzjv/ DOI: 10.17605/OSF.IO/GKZJV

## Ethical approval statement

As a systematic review of previously published research, with no primary data collection or direct involvement of human participants, ethical approval and informed consent were not required. All data included in this review were obtained from publicly accessible sources. All original research retrieved adhered to ethical guidelines.

## Notes

### Competing Interest Statement

The authors have declared no competing interest.

### Funding Statement

This study did not receive any funding

