## Supplementary file 1 for "Barriers and facilitators to implementing synchronous telehealth interventions for people with dementia - a systematic review"

**Methodological quality rating for qualitative, quantitative and mixed methods study designs using the Mixed Methods Appraisal tool (MMAT)**

| **Qualitative studies** | | | | | | | | | |
| --- | --- | --- | --- | --- | --- | --- | --- | --- | --- |
| **Study** | **Study component** | |  | **Methodological quality criteria** | | | | | |
|  | **Are there clear research questions?** | **Do the collected data allow to address the research questions?** |  | **Is the qualitative approach appropriate to answer the research question?** | **Are the qualitative data collection methods adequate to address the research question?** | **Are the findings adequately derived from the data?** | **Is the interpretation of results sufficiently substantiated by data?** | **Is there coherence between qualitative data sources, collection, analysis and interpretation?** | **Total** |
| Clark et al., 2024 | Yes | Yes |  | Yes | Yes | Yes | Yes | Yes | 5/5 |
| Cowley et al., 2022 | Yes | Yes |  | Yes | Yes | Yes | Yes | Yes | 5/5 |
| Di Lorito et al., 2022 | Yes | Yes |  | Yes | Yes | Yes | Yes | Yes | **5/5** |
| Fay et al., 2023 | Yes | Yes |  | Yes | Yes | Yes | Yes | Yes | **5/5** |
| Park et al.,2023 | Yes | Yes |  | Yes | Yes | Yes | Yes | Yes | **5/5** |
| Peri et al., 2022 | Yes | Yes |  | Yes | Yes | Yes | Yes | Yes | **5/5** |
| Perkins et al., 2022 | Yes | Yes |  | Yes | Yes | Yes | Yes | Yes | **5/5** |
| **Total** | **7/7** | **7/7** |  | **7/7** | **7/7** | **7/7** | **7/7** | **7/7** |  |

| **Randomised control trials** | | | | | | | | | |
| --- | --- | --- | --- | --- | --- | --- | --- | --- | --- |
| **Study** | **Study component** | |  | **Methodological quality criteria** | | | | | |
|  | **Are there clear research questions?** | **Do the collected data allow to address the research questions?** |  | **Are the participants representative of the target population?** | **Are measurements appropriate regarding both the outcome and intervention (or exposure)?** | **Are there complete outcome data?** | **Are the confounders accounted for in the design and analysis?** | **During the study period, is the intervention administered (or exposure occurred) as intended?** | **Total** |
| Jung et al., 2023 | Yes | Yes |  | Yes | Yes | Yes | Can't tell | Yes | 3/5 |
| Laver et al., 2022 | Yes | Yes |  | Yes | Yes | Yes | Yes | Yes | 5/5 |
| Menengiç et al., 2021 | Yes | Yes |  | Yes | Yes | Yes | No | Yes | 4/5 |
| Rhodus et al., 2023 | Yes | Yes |  | No | Yes | Yes | No | Yes | 3/5 |
| **Total** | **4/4** | **4/4** |  | **3/4** | **4/4** | **4/4** | **1/4** | **4/4** |  |

| **Quantitative non-randomised studies** | | | | | | | | | |
| --- | --- | --- | --- | --- | --- | --- | --- | --- | --- |
| **Study** | **Study component** | |  | **Methodological quality criteria** | | | | | |
|  | **Are there clear research questions?** | **Do the collected data allow to address the research questions?** |  | **Are the participants representative of the target population?** | **Are measurements appropriate regarding both the outcome and intervention (or exposure)?** | **Are there complete outcome data?** | **Are the confounders accounted for in the design and analysis?** | **During the study period, is the intervention administered (or exposure occurred) as intended?** | **Total** |
| Dial et al., 2019 | Yes | Yes |  | Yes | Yes | Yes | Yes | Can't tell | 4/5 |
| Henry et al., 2018 | Yes | Yes |  | Yes | Yes | Yes | Yes | Can't tell | 4/5 |
| Henry et al., 2019 | Yes | Yes |  | Yes | Yes | Yes | Yes | Can't tell | **4/5** |
| Lai et al., 2022 | Yes | Yes |  | Yes | Yes | No | Yes | No | **4/5** |
| Park et al., 2022 | Yes | Yes |  | Yes | Yes | Yes | No | Can't tell | **3/5** |
| Rogalski et al., 2022 | Yes | Yes |  | Yes | Yes | Yes | Yes | Yes | **5/5** |
| Sari et al., 2023 | Yes | Yes |  | Yes | Yes | Yes | Yes | Can't tell | **5/5** |
| **Total** | **7/7** | **7/7** |  | **7/7** | **7/7** | **6/7** | **6/7** | **1/7** |  |

| **Quantitative descriptive studies** | | | | | | | | | |
| --- | --- | --- | --- | --- | --- | --- | --- | --- | --- |
| **Study** | **Study component** | |  | **Methodological quality criteria** | | | | | |
|  | **Are there clear research questions?** | **Do the collected data allow to address the research questions?** |  | **Is the sampling strategy relevant to address the research question?** | **Is the sample representative of the target population?** | **Are the measurements appropriate?** | **Is the risk of nonresponse bias low?** | **Is the statistical analysis appropriate to answer the research question?** | **Total** |
| Ptomey et al., 2019 | Yes | Yes |  | Yes | Yes | Yes | Yes | Yes | 5/5 |
| Di Lorito et al., 2022 | Yes | Yes |  | Yes | Yes | Yes | No | Yes | 4/5 |
| Beeke et al., 2021 | Yes | Yes |  | Yes | Yes | Yes | Yes | Yes | **5/5** |
| **Total** | **3/3** | **3/3** |  | **3/3** | **3/3** | **3/3** | **2/3** | **3/3** |  |

| **Mixed-methods studies** | | | | | | |
| --- | --- | --- | --- | --- | --- | --- |
| **Study Component** | **Methodological quality criteria** | **Study** | | | | |
|  |  | Fanning et al., 2023 | Fredriksen-Goldsen et al., 2023 | Nicosia et al., 2023 | Paterson et al. 2023 | **Total** |
| **Screening Question** | Are there clear research questions? | Yes | Yes | Yes | Yes | **4/4** |
|  | Do the collected data allow to address the research questions? | Yes | Yes | Yes | Yes | **4/4** |
| **Qualitative** | Is the qualitative approach appropriate to answer the research question? | No | Yes | Yes | Yes | **3/4** |
|  | Are the qualitative data collection methods adequate to address the research question? | Yes | No | Yes | Yes | **3/3** |
|  | Are the findings adequately derived from the data? | Yes | Yes | Yes | Yes | **4/4** |
|  | Is the interpretation of results sufficiently substantiated by data? | Can’t tell | Can’t tell | Yes | Yes | **2/4** |
|  | Is there coherence between qualitative data sources, collection, analysis and interpretation? | Yes | Yes | Yes | Yes | **4/4** |
| **Qualitative score (/5)** | | **4** | **3** | **5** | **5** |  |
| **Quantitative randomised control trials** | Is randomization appropriately performed? | Cannot tell | n/a | n/a | n/a | **0/1** |
|  | Are the groups comparable at baseline? | Yes | n/a | n/a | n/a | **1/1** |
|  | Are there complete outcome data? | No | n/a | n/a | n/a | **0/1** |
|  | Are outcome assessors blinded to the intervention provided? | No | n/a | n/a | n/a | **0/1** |
|  | Did the participants adhere to the assigned intervention? | Cannot tell | n/a | n/a | n/a | **0/1** |
| **Quantitative Randomised control trials score (/5)** | | **1** | **n/a** | **n/a** | **n/a** | n/a |
| **Non-randomised studies** | Are the participants representative of the target population? | n/a | n/a | Yes | n/a | **1/1** |
|  | Are measurements appropriate regarding both the outcome and intervention (or exposure)? | n/a | n/a | Yes | n/a | **1/1** |
|  | Are there complete outcome data? | n/a | n/a | Yes | n/a | **1/1** |
|  | Are the confounders accounted for in the design and analysis? | n/a | n/a | Yes | n/a | **1/1** |
|  | During the study period, is the intervention administered (or exposure occurred) as intended? | n/a | n/a | Yes | n/a | **1/1** |
| **Non-randomised study scores (/5)** | | **n/a** | **n/a** | **5** | **n/a** |  |
| **Quantitative descriptive studies** | Is the sampling strategy relevant to address the research question? | n/a | Yes | n/a | Yes | **2/2** |
|  | Is the sample representative of the target population? | n/a | Yes | n/a | Yes | **2/2** |
|  | Are the measurements appropriate? | n/a | Yes | n/a | Yes | **2/2** |
|  | Is the risk of nonresponse bias low? | n/a | Yes | n/a | Yes | **2/2** |
|  | Is the statistical analysis appropriate to answer the research question? | n/a | Cannot tell | n/a | Yes | **1/2** |
| **Quantitative descriptive study score (/5)** | | **n/a** | **4** | **n/a** | **5** |  |
| **Mixed methods studies** | Is there an adequate rationale for using a mixed methods design to address the research question? | Yes | Yes | Yes | Yes | **4/4** |
|  | Are the different components of the study effectively integrated to answer the research question? | Yes | Yes | Yes | Yes | **4/4** |
|  | Are the outputs of the integration of qualitative and quantitative components adequately interpreted? | Yes | No | No | Yes | **2/4** |
|  | Are divergences and inconsistencies between quantitative and qualitative results adequately addressed? | Yes | Yes | Yes | Yes | **4/4** |
|  | 5.5. Do the different components of the study adhere to the quality criteria of each tradition of the methods involved? | Yes | No | Yes | No | **3/4** |
| **Mixed methods study score** | | **5** | **3** | **4** | **4** |  |
| **TOTALS** | | **9** | **10** | **14** | **15** |  |
