## Supplementary file 2 for "Barriers and facilitators to implementing synchronous telehealth interventions for people with dementia - a systematic review"

**Intervention characteristics**

| **Reference and country and brief name** | **Intervention objectives:** Why | **Summary of intervention:** What  (where *Goal setting* or *Tailoring* is not mentioned it was not documented in the described intervention) | **Mode of delivery:** who provided, how & where (which telehealth platform) | **Frequency:** When & How much | **Telehealth adaptation:** modification & how well | **Fidelity procedure** |
| --- | --- | --- | --- | --- | --- | --- |
| Beeke et al. (2021), UK - TeleCPT - Telehealth Communication Partner Training | To improve use of communication strategies & therefore conversation between a PwPPA & their CP | *Single component intervention*: Communication partner training  *Tailoring*: Individualised goal setting  *Goal setting:* Collaborative goal setting and goal attainment scaling. | SLT  Dyadic  Zoom | 4 weekly 1-hour sessions | Adaptation of face-to-face intervention.  Clear guidance for remote adaptation. | **✓** |
| Clark et al. (2024), Australia - Therapeutic Songwriting | To enable PLWD & their CPs to explore self-identity, relationships, and personal & shared history through songwriting. | *Single component intervention*: Guided song writing process of creating & recording lyrics and music.  *Tailoring*: Tailored to individual needs of participants | Music therapist  Dyadic and groups of dyads  Zoom | 2 x dyadic sessions + 6 group sessions (average 45 mins) | No adaptation reported. | ✘ |
| Cowley at al. (2022), Di Lorito et al. (2021) & Di Lorito et al. (2022) UK - Promoting Activity, Independence, and Stability in Early Dementia and mild cognitive impairment (PrAISED) | To promote activity and independence in people with early-stage dementia. | *Multiple component intervention*: including physical exercises, functional activities, community participation, risk enablement and environmental assessment and advice.  *Tailoring*: individually tailored programme  *Goal setting*: individualised goals based on participants preferences, needs, and aspirations. | MDT - OT, PT and support workers.  1:1 individual therapy.  Q Health | 50 x 90 min sessions over 12 months. (session duration extracted from Harwood et al, 2023) | Comprehensive remote adaptation process. | **✓** |
| Dial et al. (2019) & Henry et al. (2019), USA - PPA lexical retrieval therapies (LRT) | To maintain and generalise spoken language gains following word retrieval therapy. To compare telehealth treatment to treatment administered in person for individuals with PPA. | *Single component intervention*: lexical retrieval treatment and copy and recall therapy for home tasks.  *Tailoring*: individually tailored, functional word sets, using pictures of participants’ own objects when possible. | SLT  1:1 individual therapy  Adobe Connect | 8-16 1-hr sessions over 4-6 weeks (dosage dependent: 1 or 2 sessions per week) | Minor adaptations for the telehealth set-up. | ✘ |
| Fanning et al. (2023), USA & Canada - Virtual improvisational movement and social engagement  Intervention (IMOVE) | To positively impact QoL, brain network connectivity, motor & social-emotional functioning in people with early-stage AD & their CP | *Multiple component intervention*: combined improvisational dance, and improvisational social groups | ‘Study staff’ - not specified.  Dyadic and groups of dyads.  Zoom | Twice weekly groups and self-directed sessions for 10-16 weeks | Thorough remote adaptation & refinement process. | ✘ |
| Fay et al. (2023), Colombia - ‘TimeSlips’ storytelling  co-creation method | To positively impact personhood, QoL, and psychological well-being of Spanish-speaking PLWD in settings other than long term care. | *Single component intervention*: Facilitated group storytelling workshops. Stories are supportively elicited with a focus on creation over reminiscence.  *Tailored*: to individual interests, led by participants. Communication needs of individuals are supported. | Mixed - SLT, psychologist, 2 health academics, social worker and a project manager.  Groups.  Zoom | 32 weekly 1-hr workshops | Adaptation of a face-to-face intervention. | ✘ |
| Fredriksen-Goldsen et al., 2023, USA - Aging with Pride: Innovations in Empowerment and Action (IDEA) | To increase physical activity and problem-solving in PLWD and their CPs, who are from sexually marginalised groups | *Single component intervention*: Cognitive behavioural intervention adapted to be culturally acceptable and delivered via telehealth. | Support workers ‘Trained coaches’ (profession unspecified).  Dyadic.  Doxy.me | Not documented | Adaptation of a face-to-face intervention. | **✓** |
| Henry et al. (2018) & Dial et al. (2019), USA - Video Implemented Script Training in Aphasia (VISTA) | To improve overall intelligibility and grammaticality for trained  and untrained ‘scripts’ (rote learned functional | *Single component intervention*: script training targeting articulatory and grammatical aspects of script production, as well as memorization and conversational usage of scripted material. Video assisted speech entrainment home tasks.  *Tailoring*: Personalised and modifiable scripts to maximise achievability, functional use, and change with disease progression, adapting to the needs of participants. | SLT  1:1 individual therapy  Telehealth platform not stated. | Twice weekly (45 min to 1 h) sessions for 3m + daily home practice. | Adaptation of a face-to-face intervention.  with minor modifications for telehealth delivery | **✓** |
| Jung et al. (2023), Republic of Korea – Multimodal cognitive therapy | To investigate the effect of telehealth cognitive interventions on cognition, mood, and activities of daily living in patients with mild to moderate AD, and to compare to in-person delivery. | *Multiple component intervention*: Cognitive therapy sessions focussing on memory, attention & visuospatial function, frontal-executive function, and language. Music therapy combining improvisational singing and instrument playing. Art therapy including art appreciation, co-constructing images & memory box collage activities.  *Tailored*: Personalised art materials; music sessions tailored to individual ability and instrument preference. | MDT - therapy professions unclear.  1:1 individual & group therapy  Zoom | 32 x 30-minute ‘mixed’ sessions in 16 1-hour sessions, twice weekly for 8 weeks. | Adaptation of a face-to-face intervention.. | ✘ |
| Lai et al. (2022), Taiwan - Care and cognitive program | To provide health education, information & support to home dwelling PLWD e.g. care, assistive device and home modification options | *Single component intervention*: health promotion distance teaching. | Therapy professionals unclear.  Dyadic  Telehealth platform not stated | 1 hour, twice a week, for 8 weeks. | No adaptation reported. Designed as a telehealth study. | ✘ |
| Laver et al. (2020), Australia - Dyadic  Dementia Care Program | To improve CP skills and reduce CP stress. To maintain PLWD’s functional ability in ADLs | *Single component intervention*: Assessment and identification of home environment and key care challenges for PLWD and CPs Problem solving, education, skills building and stress management and joint working to enhance PLWD’s engagement in activities  *Tailored*: Strategies to address key care challenges are tailored to the capabilities and interests of the person with dementia, their care partner and the environment. | OT  Dyadic  Cisco Webex | 8 sessions of 1-hour over up to 16 weeks (NB. The 1st two sessions were face-to-face) | Virtual adaptation of COPE therapy (Gitlin et al, 2010). | **✓** |
| Menengiç et al. (2021), Turkey - motor-cognitive dual-task exercise intervention | To positively impact cognitive function, functional mobility, ability to perform ADLs, functional independence, depression and anxiety in PLWD. To improve CP’s well-being. | *Single component intervention*: Dual-task chair based strengthening exercise program with social elements (rapport building, singing and keeping rhythms) combined with increasing difficulty cognitive tasks.  *Tailored*: Gradual increase in session length and cognitive demand corresponding to participants’ ability. | PT  1:1 individual (CP had to be present for safety)  Zoom | 25 sessions over 6 weeks.  Session length gradually increased from 15 to 40 minutes. | No adaptation reported. | ✘ |
| Nicosia et al. (2022), USA - Preventing Loss of Independence through Exercise (PLIÉ) Mind-Body Movement Program | To support PLWD and their CPs to improve and maintain emotional well-being, physical abilities, and engagement. | *Multiple component intervention*: integrative, group movement program including functional movement,  body awareness, mindfulness, breathing, social engagement, positive psychology and music to encourage participants to move in ways that feel good to them.  *Tailored*: personalised instructions and content to facilitate social connection including participant’s interests, music preferences, and functional limitations. | Mixed - Support worker, PT, OT, yoga instructor, dance therapist, alternative therapists (Rosen & Feldenkrais).  Groups.  Zoom | 24 1-hour classes twice weekly over 12 weeks | Thorough adaptation process reported. | ✘ |
| Park et al. (2022), Park et al. (2023), USA & Canada – Remote Chair Yoga | To manage physical and psychological symptoms in socially isolated PLWD. | *Single component intervention*: Chair yoga is a safe, non-invasive, low- impact mind–body intervention for older adults with dementia, composed of physical poses, breathing and relaxation and is practised sitting in a chair or standing and using a chair.  *Tailored*: Not commented on. | PT  1:1 individual  Zoom | 16 twice weekly 1-hour sessions for 8 weeks | No adaptation process. | **✓** |
| Perkins et al. (2022) & Peri et al. (2022), Brazil, Hong Kong, India, Ireland, New Zealand & UK - Virtual Cognitive Stimulation Therapy (vCST) | To enhance cognitive functioning, social interaction, and overall well-being in individuals with mild-to-moderate dementia. | *Multiple component intervention*: including word games, social communication activities, reminiscence activities, puzzles, orientation exercises, and facilitated discussions.  *Tailored*: supportive and adaptive environment led by a skilled and trained facilitator | MDT - 'facilitators' including: clinical psychologist, OT, medics, support workers and specialist nurses.  Groups.  Zoom. | 14 x 45-60 minute weekly sessions. | Thorough adaptation process. | ✘ |
| Peterson et al. (2023), USA - CarFreeMe™ Driving Retirement Program | To assist PLWD and their CPs to manage driving retirement and find alternative transport options. | *Single component intervention*: psychoeducational coaching to help transition to driving retirement. Including education, lifestyle planning, stress management, and exploring alternative transportation options.  *Tailored*: To individual needs, circumstance, concerns and interests of participants, including the number, order and depth of sessions.  *Goal setting*: personalised driving retirement goals essential to the intervention. | MDT -nurses and clinical psychologists.  Dyadic.  Telehealth platform not stated | 4-8 1-hr sessions within a 3m timeframe | Thorough adaptation process. | **✓** |
| Ptomey et al. (2019), USA - remotely delivered exercise sessions | To positively impact moderate intensity physical activity in adults with AD and their CPs. | *Single component intervention*: Group sessions included social interaction, warm-up, moderate intensity physical activity (and cooldown. Followed by 15-minute support sessions as a dyad. | PT  Dyadic and groups  Zoom | 30-min group sessions three times weekly for 12 weeks. | No adaptation process. | ✘ |
| Rhodus et al. (2023) –  Helping older Adults cReate & Manage OccupatioNs successfully (HARMONY) | To positively impact functional occupational engagement performance and behavioural symptoms in community-residing PLWD. | *Single component intervention*: Scripted physical environment modification, caregiving and behaviour management training.  *Tailored*: To the capabilities, sensory, behavioural and caregiver needs of participants.  *Goal setting*: Individually tailored goals with occupation progression. | OT  Dyadic – with a focus on the care partner  Zoom | 6 weekly 1-hour sessions. | No adaptation process. Designed telehealth intervention. | **✓** |
| Rogalski et al. (2022), USA - Communication Bridge | To maximise participant’s QoL by facilitating communication confidence and participation in everyday situations. | *Multi component intervention*: Components include impairment activities (e.g., script training, lexical retrieval tasks), communication strategies, and disease education and support. Home tasks & check-ins facilitated by the custom web application.  *Tailored*: To individual goals and interests of individuals.  *Goal setting*: Person-centred, functional communication goals. | SLT.  Dyadic.  Custom built platform including videoconferencing, materials, goals, web-based exercises and training videos | 8 weekly 1-hr sessions + 1 pre-intervention evaluation session. Half of the dyads had 'check in' sessions. | No adaptation process. Designed telehealth intervention and platform. | **✓** |
| Sari et al. (2023), Australia & Indonesia - Telehealth Exercise Program | To improve strength, balance,  endurance and reduce falls among PLWD dementia in the community. | *Single component intervention*: Home based PT led exercise intervention including warm up, balance, resistance and walking exercises.  *Tailored*: Exercise tailored to individuals ability and to achieve moderate intensity physical activity | PT.  Dyadic.  Zoom. | 4 sessions over 12 weeks (in weeks, with 4 chock-in session + daily home practice | Adaptation of a face-to-face therapy. | ✘ |

**Key**: AD = Alzheimer’s disease, ADL=activities of daily living, CP = care-partner, FTD=frontotemporal dementia, OT = occupational therapist, PDD=Parkinson’s disease dementia, PLWD=person living with dementia, PPA = Primary progressive aphasia, PwPPA = person with PPA; PT = physiotherapist, QoL = quality of life, SLT = speech & language therapist VD=vascular dementia.
