## Supplementary file 3 for "Barriers and facilitators to implementing synchronous telehealth interventions for people with dementia - a systematic review"

**Telehealth intervention implementation facilitators identified within the TDF, COM-B and inductively derived domains, with a summary of identified facilitators**

| **TDF domain** (COM-B component) | **Frequency (number of studies identified in; maximum n=25)** |  | **Number of facilitators in this domain which pertain to the:** | | |  | **Implementation facilitators extracted from studies (number) (study)** |
| --- | --- | --- | --- | --- | --- | --- | --- |
|  |  |  | **Technology and intervention combined** | **Technology only** | **Intervention only** |  |  |
| **Environmental context and resources**  (Opportunity – physical) | 24 |  | 12 | 12 | 0 |  | - Telehealth convenience, efficiency or cost effectiveness (n = 13) (S1, S2, S3, S4, S9, S10, S11, S15, S17, S18, S19, S20, S22) - Increased access to specialists and interventions (n = 13) (S1, S3, S4, S5, S8, S9, S10, S11, S17, S19, S21, S23, S25) - Provide technical and intervention support (n = 8) (S1, S3, S6, S16, S18, S19, S23, S25) - CP support and engagement (n = 6) (S15, S17, S18, S19, S20, S24) - Equipment provision (n = 4) (S6, S9, S16, S22) - Working and optimised technology set-up prior to therapy, including in person visits to set up equipment (n = 4) (S7, S13, S14, S22) - Ensuring a detailed on-screen view of the therapists, participants, home environments and therapy resources (n = 4) (S1, S4, S18, S25) - Preparation for intervention sessions (n = 4) (S7, S15, S25) - Using own device to access telehealth (n = 1) (S19) |
| **Knowledge**  (Capability – psychological) | 20 |  | 12 | 6 | 2 |  | - Accessible telehealth platform and/or intervention adaptation guidance (e.g. manual, video) (n = 14) (S1, S3, S4, S5, S6, S7, S9, S18, S19, S20, S21, S23, S24, S25) - Training for the therapists and participants in the telehealth intervention and telehealth platforms / ‘orientation sessions’ (n = 9) (S1, S7, S9, S14, S16, S17, S18, S20, S22) - Prior knowledge & experience of telehealth or technology (n = 4) (S19, S20, S21, S22) - Accessible intervention instructions (manual, video) (n = 3) (S9, S14, S25) - Deep knowledge of participants life, needs and abilities (n = 3) (S9, S11, S14) - Therapist knowledge of dementia / experience working with people with dementia (n = 3) (S1, S4, S14) - Technical overview at the start of each session (n = 2) (S18, S20) - Regular review meetings about the technology and the intervention (n = 1) (S22) |
| **Social Influences**  (Opportunity – social) | 17 |  | 12 | 0 | 5 |  | - Opportunity to make social connections (n = 10) (S1, S2, S7, S8, S9, S12, S15, S16, S17, S18) - Peer and social support provision (n = 3) (S2, S3, S18) - In-person session(s) before telehealth intervention to build rapport and get to know participants (n = 2) (S,4, S14) - Telehealth intervention allowed (re)connection with culture, memories and relationships (n = 2) (S2, S3) - Improved dyad relationships through doing an intervention together (n = 2) (S2, S18) - Strong family and cultural values positively impacted the social nature of interventions (n = 2) (S8, S25) - Feeling of intimacy or belonging (e.g. of being in people’s homes, or part of a group) (n = 2) (S9, S18) - Dyad and group dynamic management to ensure all comfortable interacting (n = 1) (S7) |
| **Creativity**  (Inductive code) | 15 |  | 7 | 0 | 8 |  | - Creative adaptable personalised materials and input (n = 8) (S1, S6, S10, S11, S16, S20, S21, S25) - Creativity encouraged and inspired by the intervention aided engagement, fun, enjoyment and improved self-perception (i.e. intrinsic motivation) (n = 5) (S2, S3, S7, S8, S12) - Creative technology use to provide support (e.g. workarounds) and optimise the intervention (e.g. using internet resources, platform functionality) (n = 4) (S1, S2, S3, S4, S20) - Creative goal setting in-line with disease progression (n = 4) (S1, S6, S10, S11) - Pride in creative outputs of the intervention (e.g. songs, stories) (n = 2) (S2, S7) |
| **Skills**  (Capability – psychological) | 13 |  | 4 | 1 | 8 |  | - Therapist skills to optimise synchronous telehealth delivery, promote engagement and maintain the on-line space (n = 9) (S1, S2, S3, S7, S8, S15, S18, S20, S21) - Participants skill development through consistent practice and strategy uptake (n = 5) (S6, S10, S11, S15, S18) - Digital skills development through telehealth intervention (n = 2) (S7, S19) - Assessment of participant and therapist digital skills prior to interventions to see what training is required (n = 1) (S19) - Gradually increasing difficulty and duration of intervention components improves skill development (n = 1) (S15) |
| **Safety**  (Inductive code) | 12 |  | 7 | 2 | 3 |  | - Optimising the home environment for safe delivery of physical interventions (n = 5) (S3, S7, S15, S17, S18) - Remote therapist support, feedback and monitoring to manage client safety (n = 5) (S22, S25, S17, S18, S15) - In-person CP support to assist PLWD to maintain on-line, emotional and physical safety (n = 4) (S3, S15, S17, S18) - Safety training sessions and demonstration (i.e. about on-line safety, safe exercise performance) (n = 3) (S7, S9, S25) - Intervention being safe, with no adverse events (n = 3) (S17, S22, S25) - Ensuring the telehealth environment is a ‘safe space’ therapeutically and regarding on-line safety (n = 3) (S8, S20, S21) - Exercise adaptations to ensure the safe delivery via telehealth (n = 3) (S9, S15, S25) - Telehealth itself increases safety (e.g. reduces transport stress and risk, in the context of COVID-19) (n = 2) (S18, S21) - Intervention reinforces aspects of safety in terms of decision making (e.g. around driving, or care needs) (n = 1) (S21) |
| **Emotions**  (Motivation – automatic) | 12 |  | 7 | 2 | 3 |  | - Relieved burden through telehealth administration (e.g. travel, respite opportunity) (n = 6) (S7, S13, S15, S17, S18, S19) - Positive emotions (e.g. mood, raised energy levels) as a result of telehealth sessions (n = 4) (S2, S8, S19, S21) - Relieved negative emotions through telehealth intervention (n = 4) (S10, S15, S18, S21) - Feeling relaxed with telehealth set-up and intervention expectations (n = 2) (S1, S19) - CP presence improved feelings of comfort, reassurance and encouragement with telehealth administration (n = 2) (S13, S18) - Provide emotional support as part of the intervention (n = 2) (S3, S21) - Therapist enthusiasm (n = 1) (S4) |
| **Reinforcement**  Motivation – reflective | 11 |  | 2 | 0 | 9 |  | - Evidence of personal pay-off, with the application of skills, ability or decision making leading to noticeable improvements in daily life (n = 6) (S1, S2, S7, S15, S19, 21) - Reward success or progression in intervention +/- sanction of not moving on until reaching a criterion (n = 1) (S10) - Technology provides incentive and promotes intervention carryover through feedback and tracking (e.g. FitBits, video) (n = 4) (S1, S2, S22, S24) - CP and/or therapist reinforcing the benefits of therapy and providing encouragement (n = 2) (S18, S24) |
| **Beliefs about consequences**  (Motivation – reflective) | 10 |  | 2 | 3 | 5 |  | - CP / PLWD motivation to engage in the intervention through a perception it is helping (n = 5) (S14, S15, S18, S19, S20) - Belief in the effectiveness and/or ease of use of telehealth (n = 4) (S4, S13, S14, S19) - Belief in the clinical and theoretical evidence base that the intervention will be effective and lead to generalisation (n = 2) (S11, S20) - Belief in the relevance of changing strategies over time with disease progression (n = 1) (S1) - Delivering an intervention entirely via telehealth shapes users’ beliefs about its effectiveness, convenience, or accessibility (n = 1) (S14) - Testimony from previous participants (n = 1) (S20) - Taster sessions (n = 1) (S20) |
| **Beliefs about capabilities**  (Motivation – reflective) | 8 |  | 1 | 1 | 6 |  | - Participant belief in own ability and improved sense-of-self bought about through telehealth intervention with sustained impacts on life participation outcomes (n = 5) (S1, S2, S8, S15, S21, S24) - Positive perception of ability to use technology brought about through use (n = 1) (S20) - Therapists beliefs in own skills to implement a telehealth intervention and use technology (n = 1) (S1) - Therapist beliefs in CPs abilities to support telehealth (n = 1) (S5) - CPs raised awareness of partner’s strengths and resilience through the telehealth intervention (n = 1) (S8) |
| **Behavioural regulation**  (Capability – psychological) | 8 |  | 4 | 0 | 4 |  | - Practice and behaviours related to therapy targets encouraged and tracked (n = 3) (S1, S10, S11) - Self-monitoring own goals and behaviours and regularly feeding back to therapists (facilitated by telehealth systems) (n = 3) (S1, S22, S23) - Supervision to develop treatment delivery skills and improve treatment fidelity (n = 2) (S9, S14) - Participants getting into a routine of exercise or strategy use and being aware of improvements (n = 1) (S18) - Therapist reflective notes to improve treatment fidelity (n = 1) (S14) |
| **Intentions**  (Motivation – automatic) | 6 |  | 3 | 0 | 3 |  | - Intention to continue strategies, techniques or exercise due to seeing their usefulness and benefits (n = 5) (S8, S13, S18, S19, S21) - Engagement with technology due to real world need (e.g. to communicate over videoconferencing, only way to access banking) (n = 2) (S13, S16) |
| **Social/ professional role & identity**  (Motivation – reflective) | 5 |  | 1 | 1 | 3 |  | - Intervention enables participants to connect with their own personal identities and relationships (n = 2) (S2, S8) - Building the sense belonging to a group or community through the intervention (n = 2) (S8, S18) - Services / organisations able to provide the technology and staff resources to staff to be able to deliver the telehealth intervention balanced proposed cost savings (n = 1) (S20) - Self-identification as a ‘tech person’ makes it more likely to choose to do a telehealth intervention ((n = 1) S13) |
| **Goals**  (Motivation – reflective) | 5 |  | 2 | 0 | 3 |  | - Person-centred goal setting tailored to individual needs (n = 3) (S1, S3, S23) - Participants and CPs self-identifying their own achievable goals (n = 3) (S1, S21, S23) - Telehealth enabled video feedback to monitor and progress goals (n = 2) (S1, S4) - Goals set and assessed in the place they are be enacted (n = 2) (S1, S4) - Goal attainment being highly concrete, visual and contextual to a participants’ life and needs (n = 1) (S1) |
| **Memory, attention and decision processes**  (Motivation – reflective) | 4 |  | 1 | 0 | 3 |  | - Modifications to intervention sessions/activities to maximise access (e.g. sessions length and structure) (n = 3) (S15, S18, S20) - Shared decision making and joint attention inherent in intervention process (n = 1) (S8) - Real-time support (enabled by telehealth) from a therapist to attend and manage cognitive issues (n = 1) (S15) |
| **Optimism**  (Motivation – reflective) | 3 |  | 0 | 1 | 2 |  | - Telehealth intervention is fun, engaging and hopeful so participants looked forward to attending (n = 2) (S2, S8). - Therapist enthusiasm for telehealth promoted engagement (n = 1) (S4) |
| **TOTALS** | **173** |  | **77** | **28** | **68** |  |  |

**Telehealth intervention implementation barriers identified within the TDF, COM-B and inductively derived domains, with a summary of identified barriers**

| **TDF** (COM-B component) | **Frequency (number of studies identified in; maximum n=25)** |  | **Number of facilitators in this domain which pertain to the:** | | |  | **Implementation barriers extracted from studies (number) (study number(s))** |
| --- | --- | --- | --- | --- | --- | --- | --- |
|  |  |  | **Technology and intervention combined** | **Technology only** | **Intervention only** |  |  |
| **Environmental context and resources**  (Opportunity – physical) | 19 |  | 3 | 14 | 2 |  | - Limitations of telehealth medium to deliver the intervention (e.g. non-optimal view and audio, unable to pick up non-verbal cues, no physical support for exercises) (n = 12) (S1, S2, S3, S6, S7, S8, S9, S15, S16, S19, S20, S23) - No CP support, or reliance on CP increases burden (n = 10) (S4, S5, S7, S8, S13, S15, S17, S18, S19, S20) - Digital poverty: lack of access to technology (n = 8) (S1, S4, S5, S16, S17, S19, S20, S23) - Technology breakdown or access issues (e.g. due to poor functionality or design, or set up) (n = 7) (S3, S4, S5, S7, S8, S20, S23) - Telehealth funding, billing or regulation barriers (e.g. no renumeration by location or insurer, or not allowed by some services) (n = 2) (S6, S13) - Telehealth access problems due to communication, cognitive or sensory issues) (n = 4) (S1, S3, S18, S23) - Less CP peer support due to telehealth (n = 1) (S20) - Increased therapist burden using telehealth (n = 1) (S7) - Aspects of an intervention not being culturally relevant to a particular country’s context (n = 1) (S21) |
| **Knowledge**  (Capability) | 11 |  | 0 | 11 | 0 |  | - Digital literacy barrier: not knowing how to resolve technical issues (n = 7) (S1, S3, S4, S5, S17, S19, S22) - Lack of preparation (orientation, setup) (n = 3) (S3, S8, S18) - Lack of experience to deliver telehealth (n = 1) (S3) |
| **Emotions**  (Motivation – automatic) | 9 |  | 3 | 4 | 2 |  | - Stress, frustration or discomfort due to technology issues (n = 5) (S3, S4, S17, S19, S23) - Increased CP stress and burden due to participation in a dyadic telehealth intervention (n = 2) (S3, S13) - Participant anxiety due to intervention demands (n = 2) (S10, S21) - Loneliness, anxiety, and low mood are more difficult to affect or support via telehealth (n = 2) (S12, S17) |
| **Social Influences (**Motivation – reflective) | 7 |  | 4 | 3 | 0 |  | - Social preference for face-to-face therapy over telehealth (n = 3) (S4, S7, S18) - Lack of family / CP engagement (n = 2) (S13, S24) - In-person interactions which enable social connection are difficult to replicate on-line (n = 2) (S7, S20) - Insufficient time dedicated to social activities and opportunities within the telehealth intervention (n = 2) (S17, S18) |
| **Memory, attention and decision processes** (Capability – psychological) | 7 |  | 4 | 1 | 2 |  | - Cognition, memory and executive function impacting engagement in the telehealth intervention (n = 5) (S10, S11, S17, S18, S20) - Cognition, memory and executive function impacting access to telehealth platform (n = 3) (S4, S6, S17) - Memory barriers to ability to report back on practice/challenge tasks (n = 1) (S4) |
| **Safety**  (Inductive code) | 7 |  | 2 | 4 | 1 |  | - Concerns adapting physical or emotive interventions to telehealth due to no therapist in-person support, circumscribing full intervention delivery or progress (S3, S4, S8, S15, S16) (n = 5) - On-line privacy and safety concerns (n = 2) (S16, S20) - Need to have CP support to ensure safety (n = 1) (S18) |
| **Beliefs about capabilities**  (Motivational - reflective) | 5 |  | 1 | 4 | 0 |  | - Perceived low technology competence barrier: contradiction between the way participants and therapists describe their technology aptitude their actual ability (n = 4) (S1, S4, S16, S20) - Self-esteem barrier: when facing technical difficulty, participants most often assumed that they were somehow at fault (n = 1) (S16) |
| **Social / professional role & identity** (Motivation - reflective) | 3 |  | 2 | 1 | 0 |  | - Those who do not self-identify as a 'tech person', are less likely to choose to do, and engage with a telehealth intervention (n = 1) (S13) - Organisations lack local guidelines, policies or commitment to implement and sustain telehealth interventions (n = 1) (S20) - Professional identity being challenged by not looking competent with technology (n = 1) (S3) - Therapist difficulty maintaining person-centred and client focussed via telehealth (n = 1) (S3) |
| **Skills** – (Capability - psychological) | 3 |  | 1 | 1 | 1 |  | - Comprehension or communication skills barrier to benefitting from telehealth for some PLWD (n = 1) (S1) - Lack of therapist skills to navigate telehealth (n = 1) (S1) - Not introducing sufficient new material as the intervention progresses negatively impacting engagement and skill development (n = 1) (S11) - Difficulty acquiring a new technical skill for some PLWD (e.g. using Zoom functions, skill acquisition as part of therapy) (n = 1) (S25) |
| **Creativity**  (Inductive code) | 2 |  | 0 | 0 | 2 |  | - Intervention needed to be more creative and flexible (content, dosage, schedule) for individuals to maximally benefit (n = 2) (S11, S21) |
| **Reinforcement (**Motivation - automatic) | 2 |  | 1 | 0 | 1 |  | - Negative impact of non-engaged CP to due to reduced reinforcement for the PLWD to engage with telehealth or do carryover work (n = 2) (S3, S24) - Relying on CP support on-line may affect PLWD motivation and autonomy (n = 1) (S3) |
| **Goals** (Motivation reflective) | 2 |  | 0 | 2 | 0 | D | - Difficulty setting goals via telehealth, especially if required physical presence (n = 1) (S3) - Difficulty progressing goals via telehealth, especially if required physical presence (n = 1) (S4) |
| **Behavioural regulation** Capability | 2 |  | 0 | 2 | 0 |  | - Fixed habits in how to use technology impact on ability to be supported to use and problem solve with ICT (n = 2) (S16, S19) - Reduced willingness to trial new technology out of habit (n = 1) (S16) |
| **Beliefs about consequences**  (Motivation – reflective) | 2 |  | 1 | 1 | 0 |  | - Perception of telehealth groups as less effective than in-person (n = 1) (S4) - Beliefs about safety concerns (physical, on-line) impeding telehealth implementation (n = 1) (S16) |
| **Intentions**  (Motivation – automatic) | 1 |  | 0 | 0 | 1 |  | - Lack of approaches to support therapy carryover without in-person therapist oversight, support and supervision (n = 1) (S25) |
| **Optimism**  (Motivation – reflective) | 0 |  | 0 | 0 | 0 |  |  |
| **TOTALS** | **82** |  | **22** | **48** | **12** |  |  |
